# Consistent signatures in the human gut microbiome of old- and young-onset colorectal cancer

**DOI:** 10.1101/2023.11.29.23299184

**Authors:** Youwen Qin, Xin Tong, Wei-Jian Mei, Yanshuang Cheng, Yuanqiang Zou, Kai Han, Jiehai Yu, Zhuye Jie, Tao Zhang, Shida Zhu, Xin Jin, Jian Wang, Huanming Yang, Xun Xu, Huanzi Zhong, Liang Xiao, Pei-Rong Ding

**Author notes:** Corresponding authors: Youwen Qin; Pei-Rong Ding. These authors contributed equally (Youwen Qin, Xin Tong, Wei-Jian Mei).

## Abstract

The incidence of young-onset colorectal cancer (yCRC) has been increasing in recent decades, but little is known about the gut microbiome of these patients. Most studies have focused on old-onset CRC (oCRC), and it remains unclear whether CRC signatures derived from old patients are valid in young patients. To address this, we assembled the largest yCRC gut metagenomes to date from two independent cohorts and found that the CRC microbiome had limited association with age across adulthood. Differential analysis revealed that well-known CRC-associated taxa, such as *Clostridium symbiosum*, *Peptostrptococcus stomatis*, *Parvimonas micra* and *Hungatella hathewayi*, were significantly enriched (false discovery rate <0.05) in both old- and young-onset patients. Almost all oCRC-associated metagenomic pathways had directionally concordant changes in young patients. Importantly, CRC-associated virulence factors (*fadA*, *bft*) were enriched in both oCRC and yCRC compared to their respective controls. Moreover, the microbiome-based classification model had similar predication accuracy for CRC status in old- and young-onset patients, underscoring the consistency of microbial signatures across different age groups.

## Introduction

Colorectal cancer (CRC) is one of the most common non-sex-specific cancer worldwide, after lung cancer, and accounts for about one million deaths in 2020^1^. In the last few decades, the incidence of CRC has remained stable or decreased in the developed countries^2^. However, the number of young-onset CRC (yCRC, colorecta cancer diagnosed in patients under the age of 50 years) has been increasing globally^3^. The striking number of young-onset patients is a growing challenge in CRC management and global health. Despite up to 34% of yCRC have a family history of colorectal cancer^3^, the majority of cases are without clear genetic factors. The genetic background of world population is unlikely changed over the last several decades. The increasing incidence of yCRC may be attributed to changing environmental and lifestyle factors.

Among environmental factors, the gut microbiome—the microbial ecosystem residing primarily in the large intestine—has been implicated in colorectal carcinogenesis. Animal studies have pinpointed three prominent examples of gut microbial toxins in colorectal carcinogenesis^4,5^. For example, *Bacteroides fragilis* toxin promotes colon tumorigenesis by activation of the T_H_ 17 cell response^6^. Additionally, *Fusobacterium nucleatum* adhesion protein A (FadA) can bind to E-cadherin of CRC cells, activates beta-catenin signaling and regulates oncogenic responses^7^. Certain strains of *Escherichia coli* produce colibactin, a small-molecule genotoxin that can adduct to DNA and induce double-strand DNA breaks^8^. In humans, about a dozen metagenomic studies on various populations across the world have identified substantial changes in abundance of specific bacteria^9–16^. Meta-analyses of these datasets have identified globally cross-cohort microbial signatures that can predict CRC at high accuracy^13,14,17^. However, the studied cohorts have predominantly consisted of old-onset patients. The number of yCRC patients in these studies varied from 0 to 28, accumulating to 72 in total (**Supplementary Table 1**). It is uncertain whether these microbial signatures are specific to oCRC or can be generalized to yCRC.

A very recent study using 16S rRNA gene sequencing reported distinct dysbiosis in the human gut microbiome of yCRC patients^18^. Although they validated their findings in a subset of individuals using metagenomic sequencing, their main discoveries were based on 16S rRNA data. The technical limitations of this approach have limited the ability to draw definitive conclusions^19^. Further efforts based on metagenomic sequencing in larger cohort are needed to explore the gut microbial signatures in yCRC. Deep metagenomic sequencing can be leveraged to investigate strain-level diversity, providing valuable insights for experimental validation.

In this study, we generated 460 CRC metagenomes, including data from 167 yCRC patients. By integrating these data with a publicly available yCRC dataset, we identified consistent microbial signatures in both oCRC and yCRC. These signatures encompassed well-known CRC-associated taxa and virulence factors. Our strain-level analysis, focusing on three CRC-associated species (*F. nucleatum*, *B. fragilis* and *E. coli*), supported the concordance in oCRC and yCRC. We further leveraged other publicly available CRC metagenomic datasets and demonstrated that the microbiome-based predictive models had similarly high accuracy in both oCRC and yCRC patients. These results provide valuable insights into the generalizable microbial signatures of CRC and expand our understanding of CRC microbiome.

## Results

We recruited 460 CRC patients from a single hospital in Guangzhou (**Methods**). All patients were treatment naïve by the time of enrollment. Our cohort included patients with a wide age range, from 21 to 88 years old (**Figure 1a**), with 95 patients diagnosed under the age of 40 and 167 patients under the age of 50. Across all age groups, there were more male patients than female patients. 14.8% (n=68) of cancers were stage I, 32.0% (n=147) stage II, 36.1% (n=166) stage III and 17.2% (n=79) stage IV; 24.8% (n=114) were from the right hemicolon, 34.8% (n=160) left hemicolon and 40.4% (n=186) rectum; 14.6% (n=67) were with family history of CRC. There was no correlation between incidence age and sex (P=0.06), tumor stage (P=0.10), tumor location (P=0.13), and family history of CRC (P=0.3). We observed a weak correlation between body mass index (BMI) and age (Pearson correlation coefficient = 0.12, P=0.01).

**Figure 1.**
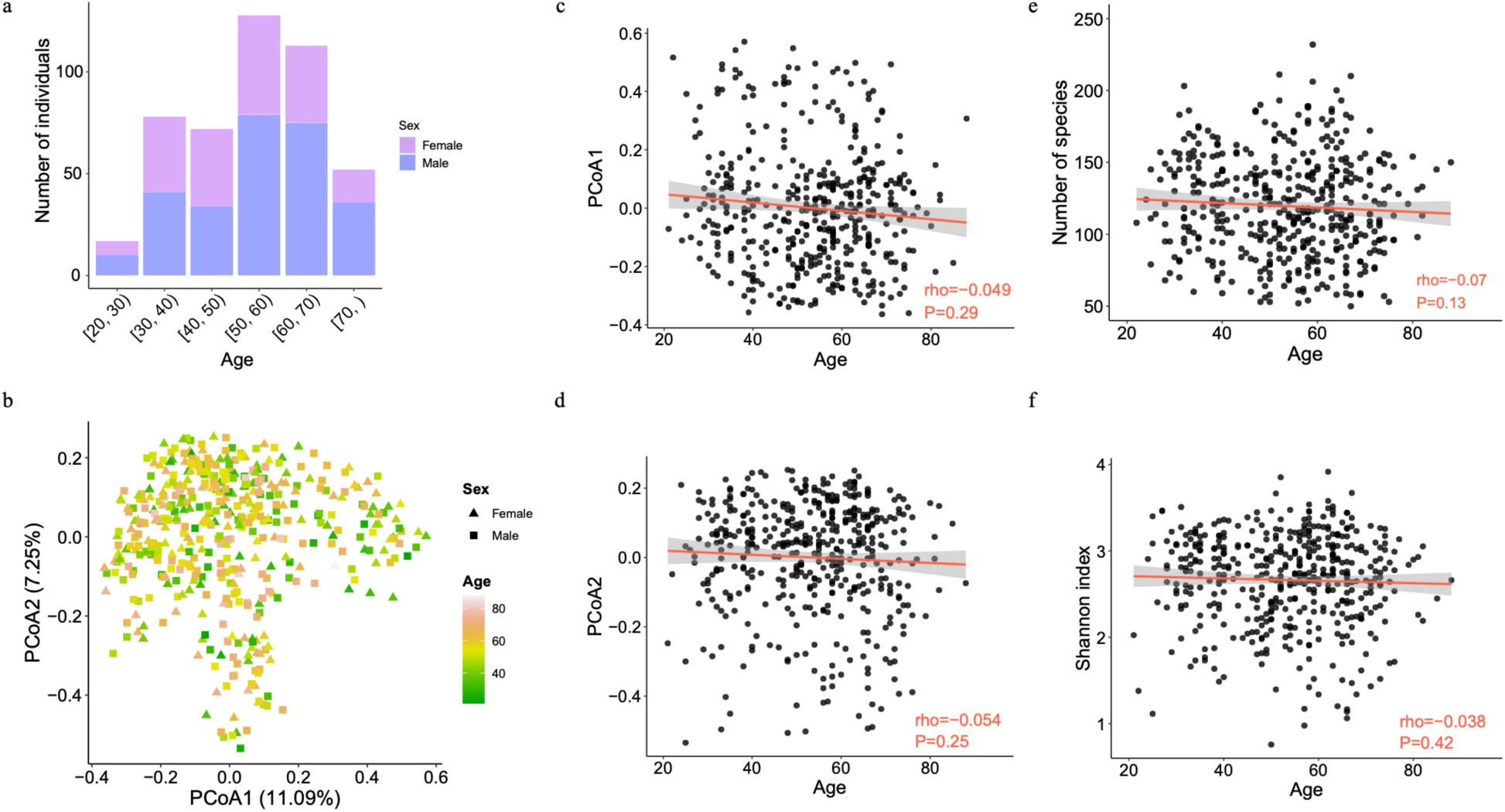
Limited association between the gut microbiome and age in CRC patients. (**a**) The number of patients in Guangzhou cohort stratified by age and sex. (**b**) Two-dimension scatter plot shows the overall pattern of samples. Principle coordinate analysis (PCoA) was performed based on the Bray-Curtis distance calculated from the abundance profile at species level. Each point represents one sample and color scale indicates age. Samples from female and male patients are in triangles and squares, respectively. Scatterplots of relationship between age and PCoA axis 1 (**c**), and PCoA axis 2 (**d**), number of species (e) and Shannon index (**f**). The solid red line was fitted by smooth function in R and the grey area is the 95% confidence interval. The Shannon index was calculated based on the abundance profile at species level.

### Limited association between the gut microbiome and age in CRC patients

The relationship between age and the gut microbiome of CRC patients was investigated using shotgun metagenomic sequencing of stool samples. We generated a total of 32,403 million paired-end high-quality reads, with an average of 70 million paired-end reads per sample (**Methods**). We found no correlation between age and alpha diversity, defined as the number of observed species and Shannon index (**Figure 1e** **& f**). The adjustment of confounders (BMI, sex, tumor location and stage and smoking) did not increase the association between alpha diversity and age. Similarly, there was no correlation between age and the first two coordinates of the principal coordinate analysis (PCoA) (**Figure 1b, c & d**). This was supported by the permutation multivariate analysis of variance (PERMANOVA) test, showing that age only explained a small fraction of microbiome variance (R^2^=0.003, P=0.15).

To identify specific taxa associated with age, we tested the correlation between age and species abundance (**Methods**). We only found that the abundance of four species, namely *Prevotella stercorea*, *Bifidobacterium dentium*, *Prevotella copri* and *Prevotella bivia*, were significantly correlated with age (false discovery rate (FDR) < 0.05, **Supplementary Table 2**). The associations of *Prevotella* species were negative, while that of *B. dentium* was positive. *B. dentium* and *P. bivia’s* associations with age were independent of body mass index (BMI), sex, tumor location and stage, family history of CRC and smoking. Although *Prevotella* species are commonly found in the human gut microbiota and have been linked to dietary habits, their relative abundances were age-dependent and dropped from adulthood to old age^20^. *B. dentium*, which is commonly found in the human oral microbiome^21^, had increased abundance and prevalence in the gut with age^22^. While two dozen of other bacterial species have been identified as age-associated^23^, their associations with age in CRC patients were weak, suggesting that CRC status may outperform age in shaping the gut microbiome.

As CRC is one of the most studied traits in gut microbiome research, a collection of CRC-associated taxa has been robustly identified in previous studies. We obtained a list of 118 CRC associated taxa from gutMDisorder^24^ (**Methods**). Among them, 24/25 taxa reported in at least two studies were present in our cohort with prevalence rates from 15.43% to 90.65% (average 58.41%), with 16 taxa presented in over half patients. Importantly, only *P. copri* abundance was correlated with age, but its correlation was not significant after adjustment for confounders (**Supplementary Table 2**).

Additionally, we validated our findings in a recently published yCRC cohort (Fudan cohort)^18^. Age was given as a binary variable, old or young with the cutoff of 50, in the Fudan cohort. Consistently, we separated our Guangzhou patients into old (age>50) and young (age<50) groups. We only found nine species with differential abundance (P<0.05) between old and young groups in both cohorts (**Supplementary Table 3**). Four of them (*P. stercorea*, *B.dentium*, *P. copri* and *P. bivia*) were mentioned above. Although the other five species included previously reported CRC-associated microbe *Alistipes indistinctus*^10,15^, and CRC-depleted microbe *Eubacterum rectale*^10^ and *Faecalibacterium prausnitzii*^10^, none passed the multiple testing adjustment significance threshold (FDR<0.05). Taken together, there was limited (if any) association between the known CRC-associated taxa and age.

### Bacterial species associated with oCRC and yCRC

To investigate the gut microbiome changes in oCRC and yCRC patients, we compared them to age-matched controls. We reanalyzed the stool microbiome data from the published yCRC cohort (Fudan cohort)^18^ (**Methods**) and integrated with our Guangzhou cohort. In accordance with Yang et al^18^, the yCRC was defined as age under 50 years old, and the others were oCRC. A PCoA based on species level abundance showed that disease effect surpassed the batch effect (**Figure 2a, b & c**). The Guangzhou patients had similar distributions in PCoA1 and PCoA2 with patients in the Fudan cohort, and lower median values than the controls. Furthermore, the CRC status explained slightly higher variance than study effect (R^2^= 0.00408 and 0.00376, PERMANOVA). Therefore, the batch effect in Guangzhou and Fudan cohorts was limited.

**Figure 2.**
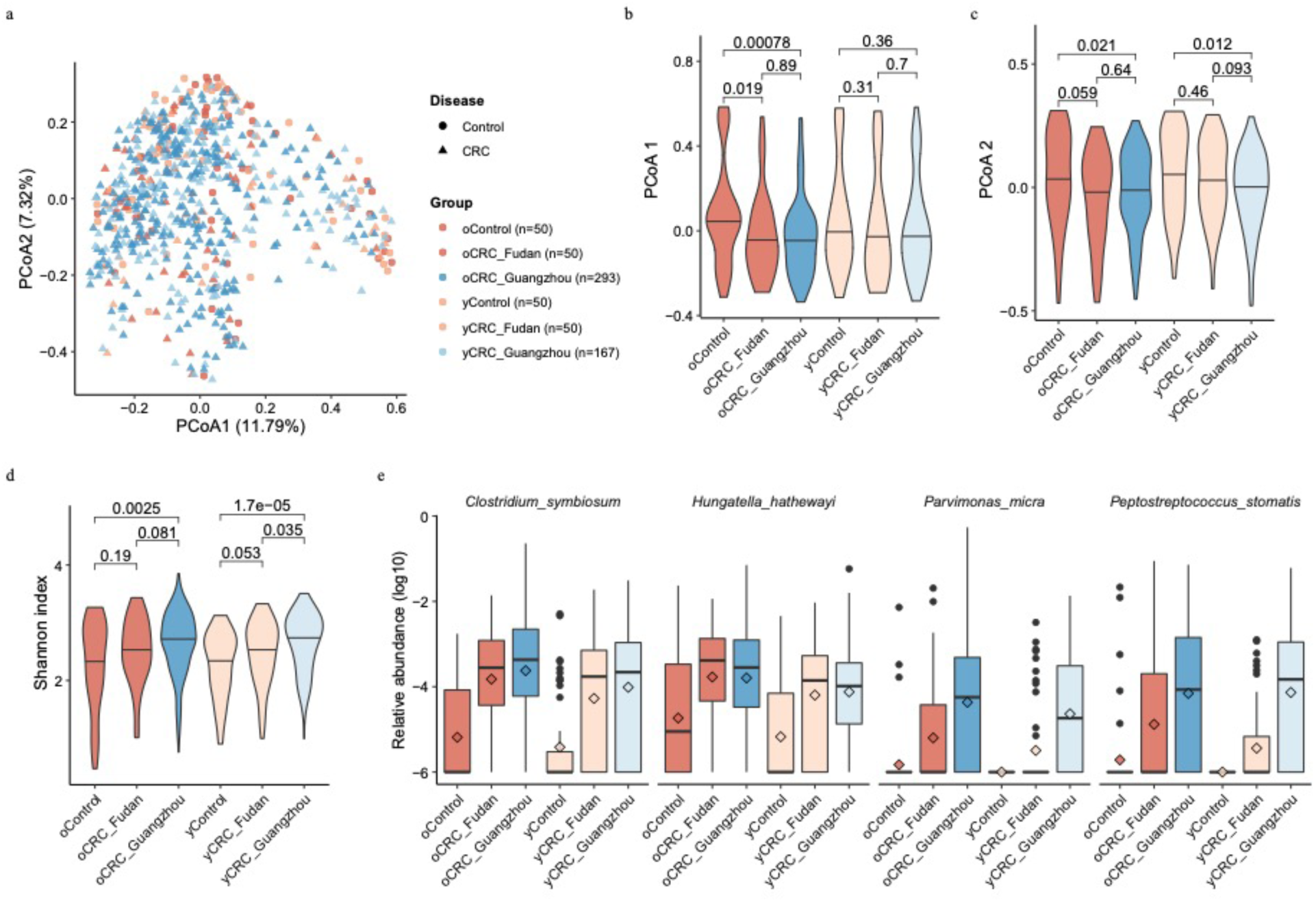
Consistent changes of CRC-associated microbes in old- and young-onset patients in two independent cohorts. (**a**) Two-dimension scatter plot shows the overall distribution of Fudan and Guangzhou samples. Principle coordinate analysis (PCoA) was performed based on the Bray-Curtis distance calculated from the abundance profile at species level. Each point represents one sample. Samples from Fudan and Guangzhou cohorts are in red and blue, respectively. Circles are control samples, while triangles are CRC samples. Violin plots show values of PCoA axis 1 (**b**), PCoA axis 2 (**c**), and Shannon index (**d**) across different groups. The thick horizon line indicates the 50% percentile. P values on the top were calculated by Wilcoxon rank-sum test. (**e**) Four well-known CRC-enriched species significantly enriched in both oCRC and yCRC patients at false discovery rate (FDR) adjusted P<0.05. The relative abundance is in log10 scale and zeros were replaced by a small value. The box plots show the median (thick line), interquartile range (box limits), 1.5× the interquartile range span (whiskers) and outliers (dots). Diamond shape indicates the mean abundance.

Previous studies have suggested that the microbiome of CRC patients had a higher alpha diversity than controls, possibly due to the expansion of typically oral microbes in addition to the baseline gut microbiome^13,14,17^. We confirmed this finding in the Fudan cohort, where oCRC and yCRC patients had a higher Shannon index than their controls (**Figure 2d**). Guangzhou patients also had a higher Shannon index than the Fudan controls, supporting the increased diversity in CRC patients.

To identify taxa that are differentially abundant in CRC patients, we conducted two sets testing. The first set of testing was conducted only on the Fudan cohort, while the second set was conducted on the Guangzhou patients and Fudan controls (**Methods**). Our analysis revealed four species (*Clostridium symbiosum*, *Peptostrptococcus stomatis*, *Parvimonas micra* and *Hungatella hathewayi*) that were consistently enriched (FDR adjusted P<0.05) in oCRC and yCRC patients in both cohorts compared to Fudan controls (**Figure 2e**, **Supplementary Table 4**). These four species are well-known CRC-associated biomarkers^13,14^ and were not associated with age (**Supplementary Table 3**). *C. symbiosum*, for instance, was first reported by a qPCR study^25^ and confirmed in a meta-analysis study which integrated five shotgun metagenomic studies^13^. *P. micra* and *P. stomatis* were among the most important features in CRC classifiers built on stool microbiome data^13^.

We also found six other microbial species that were differentially abundant (FDR adjusted P<0.05) in oCRC and showed similar trends in yCRC in both cohorts (**Supplementary Figure 1, Supplementary Table 4**). Among these six taxa, *Eggerthella lenta*, *Erysipelatoclostridium ramosium* and *Flavonifractor plautii* were enriched in CRC groups and have been previously reported as biomarkers for CRC^10,18^. In particular, *F. plautii* was identified as a biomarker for yCRC in a study by Yang et al^18^. Two known bacteria *Eubacterium rectale* and *Ruminococcus bicirculans*, as well as a metagenomically assembled taxon *Eubacterium sp. CAG38* were depleted in CRC microbiome. *E. rectale* is one of the most prevalent human gut bacteria^26^ and was reproducibly reported with decreased abundance in CRC patients compared to healthy controls^9,10^.

On the other hand, three of the four taxa (*Alistipes indistinctus*, *Clostridium aldenense*, *Eisenbergiella tayi* and *Fusobacterium sp oral taxon 370*) enriched in yCRC showed similar trends in oCRC as well (**Supplementary Figure 2, Supplementary Table 4**). *Fusobacterium sp oral taxon 370* is one of the typical oral bacteria linked to CRC in old-onset patients^14^. While *A. indistinctus* was at a low abundance in the human gut microbiome, it was involved in CRC carcinogenesis and treatment response^27^. We specifically analyzed two taxa, *B. fragilis*^6^ and *F. nucleatum*^7^, with proposed carcinogenesis mechanism. Although the *B. fragilis* abundance was higher in CRC than control, the significance was lost after multiple hypothesis adjustment (**Supplementary Figure S3**). *F. nucleatum*’s prevalence was surprisingly low in the Fudan cohort, while its abundance had a similar distribution in oCRC and yCRC of the Guangzhou cohort. Additionally, at the significant level of nominal P<0.05, 23 of 24 species passing the threshold had directionally consistent changes in old- and young-onset patients compared with their controls **(Supplementary Table 4**). Overall, our findings indicate that most of the CRC-associated taxa showed concordant changes in both oCRC and yCRC microbiomes compared to their controls.

### Strain-level analysis of *F. nucleatum*, *B. fragilis* and *E. coli*

Our deep metagenomics sequencing data allowed us to investigate the strain-level insights into CRC-associated species. Given the inherent challenges of strain-level, we focused on three CRC-associated species *F. nucleatum* (*Fn*)*, B. fragilis* (*Bf*) and *E. coli* (*Ec*) in the Guangzhou cohort. We used StrainPhlAn 3^17^ to construct the phylogenetic tree and used inStrain^28^ to examine the genome-wide sequence diversity. (**Methods**).

*Fn* was identified in 63 samples according to its marker-genes and the corresponding phylogenetic tree showed no correlation with patient age, tumor stage and location (**Supplementary Figure S4a**). No significant difference was observed in the *Fn* prevalence between oCRC and yCRC. Genome-wide sequence analysis revealed similarly high population-level average nucleotide identity (popANI)^28^ values to the *Fn* reference genome in oCRC and yCRC metagenomes, with no significant difference (**Supplementary Figure S4b**). We also evaluated the nucleotide diversity, an indicator of strain diversification. Our analysis revealed no association between *Fn* nucleotide diversity and patient age, tumor stage or location (**Supplementary Figure S4c-e**). A study reported that *F. animalis* (*Fa*, also known as *Fn subsp. animalis*) had higher abundance and prevalence than *Fn* in tumor samples^29^. In line with this finding, we found *Fa* in more samples with higher coverage than *Fn* in our cohort (**Supplementary Figure S4f**). *Fa* prevalence, popANI and nucleotide diversity values showed no difference in oCRC and yCRC (**Supplementary Figure S5**). Taken together, the analysis of *Fn* and *Fa* distribution and diversity revealed no distinctions between oCRC and yCRC. However, we noted a higher nucleotide diversity of *Fa* in colon compared to rectum tumors (**Supplementary Figure S5**). A similar trend was observed for *Fn*, albeit with lower significance due to smaller sample size. This observation suggests that *Fn* and *Fa* exhibited increased diversity in patients with colon tumors.

For *Bf*, we identified two distinct phylogenetic clusters (**Figure S6a**). Cluster 1 (N=267) was dominated by samples with metagenome-assembled genomes (MAGs) annotated to strain NCTC9343 (average nucleotide identity (ANI) >95%), while cluster 2 (N=67) was dominated by samples with MAGs annotated to strain Q1F2 (**Methods**). The distribution of oCRC and yCRC was not different in the two clusters. The phylogenetic tree revealed no correlation with patient age, tumor stage or location, within or between clusters (**Supplementary Figure S6a**). Genome-level analysis demonstrated high popANI values to the reference genomes for both strains in oCRC and yCRC (**Supplementary Figure S6b**). While the nucleotide diversity of cluster 1 was not associated with age, the nucleotide diversity of cluster 2 was higher in yCRC than oCRC (P=0.02, **Supplementary Figure S6c**). This suggests that *Bf* strain in cluster 2 samples may be under stronger selection pressure in yCRC patients.

For E. coli, our analysis identified only one strain cluster (ANI >95% to strain ATCC 11775) in 317 (69%) samples, with no significant difference in prevalence between oCRC and yCRC. The marker-gene based phylogenetic tree analysis revealed no correlation with patient age, tumor stage or location (**Supplementary Figure S7a**). Genome-level analysis indicated a similar popANI value to the reference genome in oCRC and yCRC (**Supplementary Figure S7b)**. Nucleotide diversity analysis showed no association with patient age, tumor stage or location (**Supplementary Figure S7c-e**).

### Functional metagenomic signatures for oCRC and yCRC

Unlike 16S rRNA gene amplicon data, metagenomes allow us to access the functional capacity of the gut microbiome. In our Guangzhou cohort, age did not have a significant association with the top two axis of PCoA, calculated from the microbial pathway profile (**Supplementary** Figure 8). In PERMANOVA, age explained a small and non-significant amount of overall variance of the microbial pathway variation (R^2^=0.005, P=0.13). To identify specific microbial pathway associated with age, we tested the association between age and abundances of metaCyc pathways (**Methods**). Only one metaCyc pathway, PWY-6608: guanosine nucleotides degradation III, was associated with age at P<0.01 with and without adjustment for confounding factors (**Supplementary Table 5**).

To identify functional signatures associated with oCRC and yCRC, we integrated the functional pathway profiles of our Guangzhou cohort and the published Fudan cohort. PCoA showed that Guangzhou patients were closer to Fudan patients than controls (**Supplementary** Figure 9). In addition, CRC status explained higher variance than the study effect (R^2^=0.014 vs. 0.006, PERMANOVA). We then conducted differential analysis in metaCyc pathways (**Methods**). Out of the 435 tested metaCyc pathways, 69 had differential abundance between oCRC and oControl in both cohorts (FDR adjusted P<0.05, **Supplementary Table 6**). Among these, while only one pathway, PWY-7316: dTDP-N-acetylviosamine biosynthesis, was differential in yCRC and their controls at the same significant level, the majority (60/69) had concordant enrichment direction in yCRC.

We next examined the well-known CRC-associated microbial *cutC* gene, which encodes choline trimethylamine-lyase responsible for production of the disease-associated trimethylamine (TMA). In total, 113 UniRef90 gene families annotated as *cutC* orthologs were detected in at least one sample in Guangzhou or Fudan cohorts. Of these, 8 *cutC* orthologs presented in more than half of Guangzhou patients had concordant enriched direction in oCRC and yCRC compared to their controls (**Supplementary Table 7**). Remarkably, two *cutC* orthologs with represented sequences from *Bacteroides* species, including *Bf*), reached a statistically significant level of nominal P<0.05. The sum abundance of *cutC* gene families was higher in oCRC and yCRC compared to their controls (**Supplementary** Figure 10).

We accessed CRC-associated virulence factors and toxins, focusing on *fadA* (encodes *Fn* adhesion protein A)^7^, *bft* (encodes *Bf* enterotoxin)^5^, the *pks* genomic island (encodes colibactin in some *Ec* strains)^8^ and the *bai* operon (encodes enzymes for the conversion of primary to secondary bile acids in *Clostridium* species)^30^ (**Methods**). *fadA* exhibited significant enrichment in both oCRC and yCRC compared to their respective controls (**Figure 3a**). In the strain-level analysis, we identified *Fn* and *Fa* in our samples. Here we further explored *fadA* abundance in the context of *Fn* and *Fa*. *fadA* abundance was not different in samples with *Fn* or *Fa* (**Figure 3b**). Samples with both *Fn* and *Fa* had the highest *fadA* average abundance, while samples without any strain exhibited the lowest *fadA* average abundance. For *bft*, we observed an enrichment trend in CRC compared to controls (**Figure 3a**). Notably, both *bft* abundance and prevalence were differentiated in the two phylogenetic clusters defined in the strain-level analysis section (**Figure 3c**). The *pks* abundance exhibited variability between cohorts, being higher in CRC than controls in the Fudan cohort but lower in the Guangzhou CRC than controls (**Figure 3a**). Strain-level analysis identified strain *Ec* NCTC9343 was the dominant strain in the Guangzhou cohort, and the reference genome of this strain did not contain the *pks* island. We explored the correlation between *Ec* and *pks* abundances, finding a positive correlation in the Guangzhou cohort but no correlation in the Fudan cohort (**Figure 3d**). This discrepancy suggests a potential difference in the ecological context between populations. For *bai*, its abundance was significantly higher in yCRC than the respective controls (**Figure 3a**). In the Fudan cohort, oCRC and their controls had similar level of *bai*, which were higher than those in young controls. This finding aligns with reports of elevated bile acid metabolism in elderly people^31^, indicating that ageing may influence the association between *bai* and CRC in the elderly. In summary, CRC-associated virulence factors (*fadA*, *bft*) were enriched in both oCRC and yCRC.

**Figure 3.**
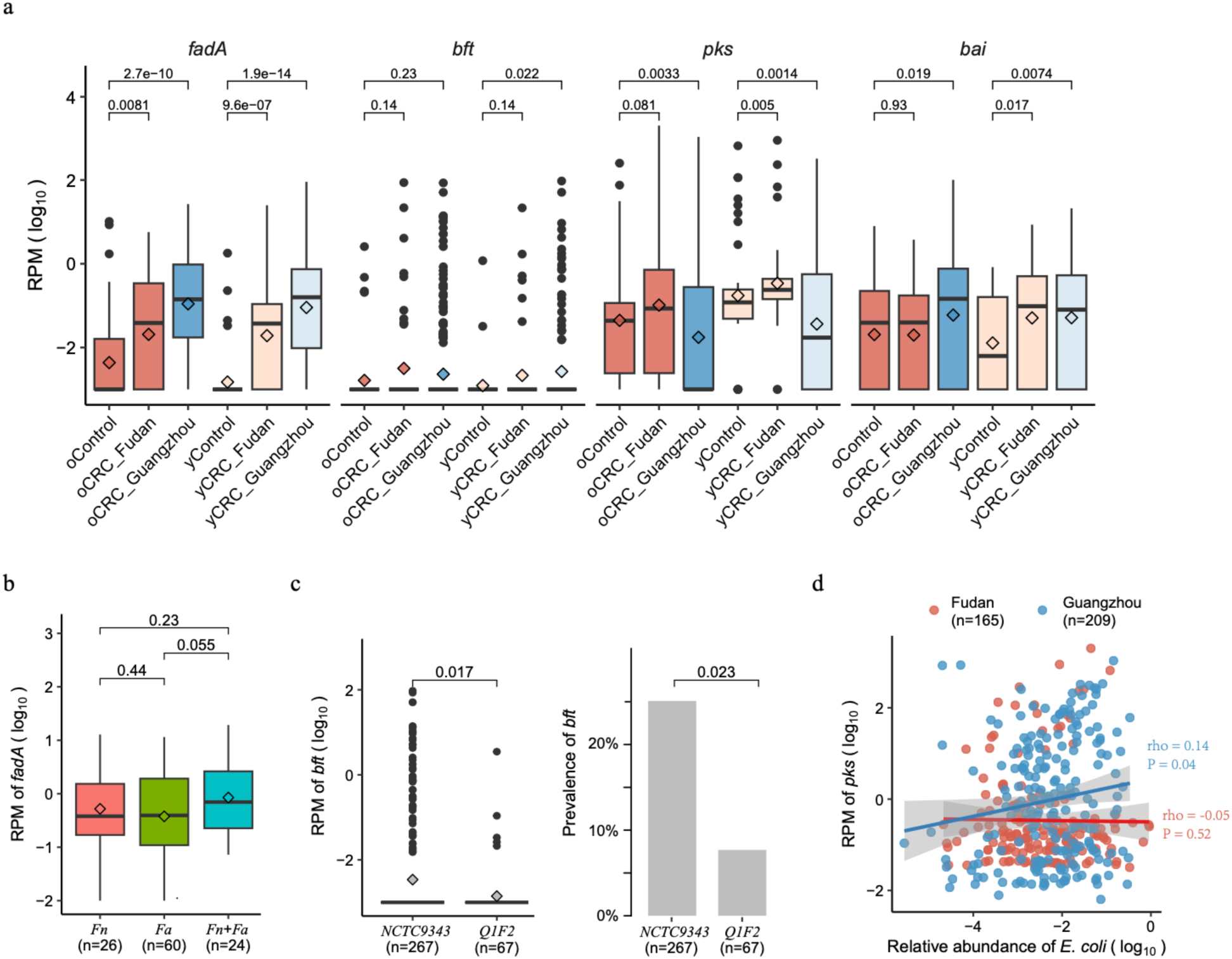
Enrichments of CRC-associated virulence factors and toxins in old- and young-onset patients in two independent cohorts. (a) Normalized log abundances of CRC-associated virulence factors in different groups. RPM means reads per million mapped reads, see **Methods** for gene quantification. *fadA* encodes *F. nucleatum* adhesion protein A; *bft* encodes *B. fragilis* enterotoxin; the *pks* genomic island encodes enzymes to produce genotoxic colibactin (in *E. coli*); the *bai* operon encodes bile acid-converting enzymes (present in some Clostridiales species). (**b**) Normalized log abundances of *fadA* stratified by the presence of *F. nucleatum* (*Fn*) and *F. animilis* (*Fa*). Presence was defined as genome breadth >0.1 and coverage >0.1. (**c**) Normalized log abundances and prevalence of *bft* stratified by *B. fragilis* strain clusters. Cluster assignment was conducted based on marker genes and genome-wide sequence analysis (**Methods**). (**d**) Correlation between normalized log abundances of *pks* and *E. coli*. The correlation coefficient was calculated using the Spearman method. The boxplot conventions are consistent with the description in Figure 2.

### Similar prediction accuracy of CRC status in old- and young-onset patients

Several studies have demonstrated the potential of tailoring the gut microbiome for predicting CRC status^13,14,17^. To access the transferability of classifiers across different patient groups, we employed the random forest algorithm to train machine learning models separately on oCRC and yCRC patients (**Methods**). Our results revealed promising cross-application performance. The model trained on Fudan oCRC patients exhibited robust predictive capability for Fudan yCRC patients, achieving an area under receiver operator curve (AUROC) of 0.7688, only slightly lower than the cross-validated AUROC of 0.8127 (**Figure 4a**). Similarly, the model trained on Fudan yCRC patients performed similarly to the cross-validation on oCRC patients, with an AUROC of 0.7548 and 0.7671, respectively (**Figure 4a**). We extended our evaluation to the Guangzhou cohort. The oCRC model demonstrated a high recall rate of 0.7952 when predicting CRC status in Guangzhou oCRC patients, and surprisingly, it outperformed the yCRC model in predicting CRC status in Guangzhou yCRC patients (0.7485 vs. 0.5629, **Figure 4b**).

**Figure 4.**
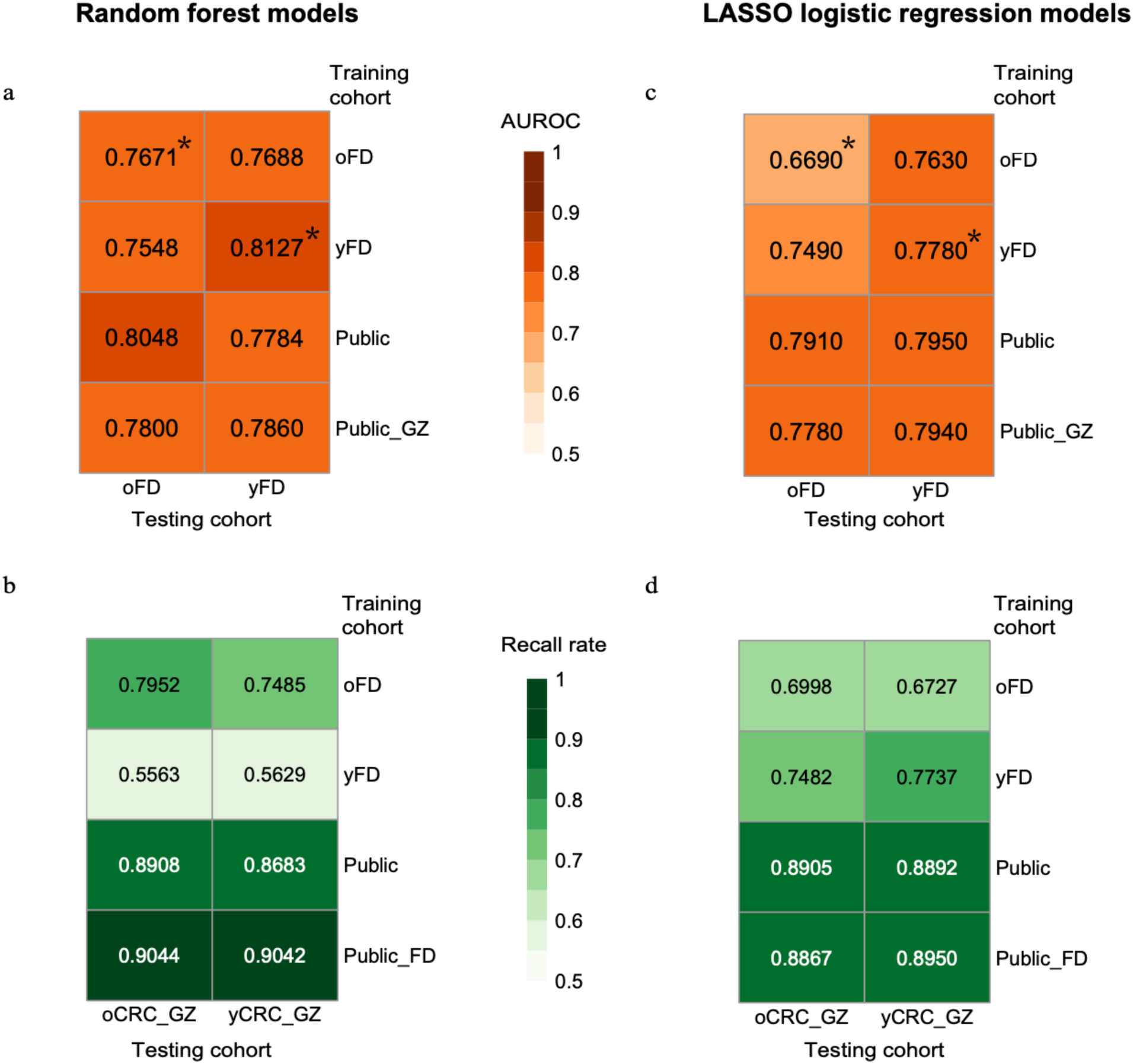
Prediction accuracy of CRC status in old- and young-onset patients. Prediction performance on oCRC and yCRC in Fudan cohort and Guangzhou cohort using models trained on species-level abundances from different datasets. Models were trained on two different methods: random forest and LASSO logistic regression. The numerical values are the area under receiver operator curve (AUROC) for (**a**) and (**c**), and recall rate for (**b**) and (**d**). Asterisks denote the values averaged over 100 times of ten-fold cross-validation. Abbreviations: oFD means the 100 metagenomes of Fudan oCRC and oControl; yFD means the 100 metagenomes of Fudan yCRC and yControl; Public means 1,262 public metagenomes; Public_GZ means 1,262 public metagenomes plus 460 Guangzhou metagenomes; Public_FD means 1,262 public metagenomes plus 200 Fudan metagenomes.

To overcome the limitation of a small sample size in the Fudan cohort, we incorporated the publicly available dataset that consisted of 600 CRCs and 662 controls, which included only 72 patients diagnosed under the age of 50^9–16^. The model trained on this expanded dataset predicted CRC status slightly better in Fudan oCRC patients compared to yCRC patients (AUROC=0.8048 vs. 0.7784, **Figure 4a**). Similarly, the model predicted CRC status in the Guangzhou cohort with a slightly higher recall rate in oCRC than yCRC patients (0.8908 vs. 0.8683, **Figure 4b**). When we included Guangzhou CRC patients in the training data, the resulting model had similar prediction accuracy for oCRC and yCRC patients in the Fudan cohort (AUROC=0.7800 and 0.7860, **Figure 4a**). The model trained on the public and Fudan datasets also showed similar performance on oCRC and yCRC patients in the Guangzhou cohort (recall rate=0.9044 and 0.9042). In summary, our results suggest that the microbiome-based classifiers can accurately predict CRC status in both old- and young-onset patients.

Additionally, we trained the random forest models using metagenomic pathway profiles to predict CRC status (**Methods**). The overall performance of the pathway-based model, as measured by the AUROC and recall rate, was lower than that of the species-based model (**Supplementary** Figure 11). This aligns with previous studies reporting that metagenomic pathway-based CRC prediction models tend to exhibit relatively poorer performance compared to species-based models^13,17^.

Finally, we replicated our machine learning experiments using another method, least absolute shrinkage and selection operator (LASSO) logistic regression. Consistent with the findings from the random forest approach, the LASSO models built on species profiles performed better than the pathway profiles (**Figure 4c-d** **& Supplementary Figure 11c-d**). Importantly, the performance of LASSO models on oCRC and yCRC was similar, suggesting the models’ transferability across different age groups.

## Discussion

The CRC microbiome has been extensively studied in old-onset patients, but less is known in young-onset patients. By integrating public data with our own data, we assembled the largest metagenome dataset of young-onset CRC patients to date. Our results revealed that the gut microbiome changes associated with CRC were similar in both old- and young-onset patients in two independent cohorts. We observed the enrichment of key CRC-associated bacteria, including *P. micra*, *P. stomatis*, *C. symbiosum* and *H. hathewayi*, in both patient groups. CRC-associated virulence factors (*fadA*, *bft*) were enriched in both old- and young-onset CRC compared to their respective controls. Our strain-level analysis reinforced the consistency of CRC-associated microbial signatures in old- and young-onset CRC. Additionally, the similar prediction accuracy of our microbiome-based models for CRC status in both old- and young-onset patients underscores the consistency of microbial signatures across different age groups.

In contrast to the widespread assumption that intestinal dysbiosis is generally associated with decreased alpha-diversity, the gut microbiome in CRC exhibits higher richness than controls^13,14^. Our findings indicated that the increased alpha-diversity was not only observed in old-onset patients but also in young-onset patients in both the Guangzhou and Fudan cohorts. However, in the original study of the Fudan cohort, they reported a reduction in alpha-diversity in old-onset CRC but an increase in young-onset CRC based on 16S rRNA gene amplicon sequencing^18^. The observed discrepancy may stem from the methodological differences between metagenomic and 16S rRNA gene amplicon data. The robustness of our metagenomic approach in capturing microbial richness highlights the importance of considering sequencing methodologies in microbiome research.

Discrepancy was also found in individual taxa. The Fudan study identified *F. plautii* as an important bacterial species in yCRC, but not in oCRC^18^. We confirmed its enrichment in young-onset CRC patients in our Guangzhou samples. Whereas the abundance of *F. plautii* was also increased in oCRC in Guangzhou and Fudan cohorts according to metagenome data. Moreover, the four disease-enriched species (*P. micra*, *P. stomatis* and *C. symbiosum* and *H. hathewayi*) were not distinguishable in 16S rRNA data between CRC and controls in Fudan study^18^. These discrepancies may be accounted by the better resolution of species level profiling offered by metagenomic data^32^. Notably, *F. nucleatum* was not among the CRC-associated features in Fudan study^18^. Through genome-wide sequence analysis, we confirmed that *F. nucleatum* can only be detected (with breadth >0.1 and coverage >0.1) in two Fudan samples. The low prevalence of this species did not provide a sufficient sample size for rigorous statistical testing.

Accumulating evidence has shown that insights into microbial strain-level are essential for understanding disease associated commensal bacteria^5^. Our strain-level analysis of the deep sequencing data found two closely related *Fusobacterium* species *F. nucleatum* and *F. animalis* in the stool metagenome of CRC. This finding confirmed a previous study that reported *F. animalis* as more abundant than *F. nucleatum* in CRC tumor biopsies^29^. The abundance of *fadA* was equally high in samples containing either *F. nucleatum* or *F. animalis*, suggesting that these two species may be pathogenic. More importantly, we found two *B. fragilis* strain clusters in CRC. The abundance and prevalence of enterotoxin gene *bft* was higher in cluster with strains closely related to NCTC 9343 than cluster with strain closely related to Q1F2. This example supports existing literature on the complex roles of *B. fragilis* in human health and disease^5^. However, further studies are required to determine the identity of these strains.

Our metagenomic functional analysis revealed an age-independent enrichment of the N-acetylviosamine biosynthesis pathway in the CRC microbiome. Given that N-acetylviosamine is a component of the O-antigen, which is part of the lipopolysaccharide (LPS)^33^, our findings align with a recent study indicating LPS enrichment in blood samples from CRC patients^34^. As LPS is a prevalent product in the human gut microbiome, capable of activating Toll-like receptor 4 and inducing immune responses and inflammation^35^, our results support the immune and inflammation hypothesis of CRC carcinogenesis^4^.

While our study contributes valuable insights into the age-independent changes in the gut microbiome associated with CRC, it is crucial to acknowledge certain limitations. Given the incidence rate of young-onset CRC is about an order of magnitude lower than old-onset CRC^36^, the number of young-onset patients in our study was smaller than that of old-onset patients. Despite this limitation, our investigation remains the most extensive analysis of young-onset CRC patients to date. Tumor molecular characteristics, such as mismatch repair and BRAF status, are essential for studying host-microbe interactions. Although our CRC sample size was relatively large, the number of patients with mutations in molecular biomarkers was limited (**Supplementary Table S8**). Consequently, we were unable to draw robust conclusions regarding the association between molecular biomarkers and microbiome features. Moreover, we cannot definitively establish whether the CRC enriched microbes are merely associated with the disease or have causal roles in carcinogenesis. Despite age-related changes in the gut microbiome observed in the general population^23^, our results unveiled convergent changes in gut microbiome in old- and young-onset CRC patients, regardless of age. This implicates that the tumor status may drive the alteration of microbial ecosystem in the gut, surpassing the age-related changes. It is important to note the limitations stemming from the absence of dietary information in our study. Dietary factors play an important role in shaping the gut microbiome. Given the complexity of Chinese cooking culture (characterized by diverse and complex dishes)^37^, obtaining precise and reliable dietary information was out of the scope of this study. The absence of dietary information in our study emphasizes the need for future investigations to explore the intricate interplay between diet, convergent changes in the gut microbiome, and their potential roles in promoting carcinogenesis.

Microbiome-based models have been explored and strongly support the promise of non-invasive CRC diagnostics^13,14^. Our study expanded this promise by demonstrating that the model could be applied across a broad age range. Fecal sample has been recognized as an ideal source for non-invasive CRC screening. Currently, the main large-scale implemented CRC screening tests include the fecal immunochemical test (FIT) and the fecal DNA methylation test. However, both methods lack an optimal threshold for young and old populations, as the fecal hemoglobin concentration varies with age and sex^38^ and DNA methylation changes are more likely absent in young patients than old patients^39^. The similar prediction accuracy of the gut microbiome-based prediction model shown in our study may facilitate the generalization of CRC screening to all adulthood.

In conclusion, our study highlights the age-independent signatures in the gut microbiome of CRC. The identified microbial patterns emphasize the potential of microbiome-based models for non-invasive CRC diagnostics across a diverse age group. Our findings also demonstrate an example of investigating disease-associated microbial signatures at the strain-level, contributing to a more nuanced understanding of the intricate relationships between the microbiome and CRC.

## Materials and Methods

### Study cohorts and data

#### Guangzhou cohort

All patients were recruited from Sun Yat-sen University Cancer Center in Guangzhou in accordance with the study protocol approved by the Ethics Committee of Sun Yat-sen University Cancer Center (B2019-214-X02). Informed consent was obtained from every patient. The inclusion criteria were newly diagnosed pathologically proven locally advanced rectal adenocarcinoma. Patients with any of previous tumor history, tumor treatment history, antibiotics and/or probiotics treatment within one-month were removed from the study. Clinical data was obtained from regular medical checkups and questionaries. The summary metadata information of recruited patients was given in **Supplementary Table 8**.

Stool samples were collected by patients following the manufacturer’s instructions either at home or in the hospital^40^. Samples were transferred to the laboratory and stored at -80 °C within 7 days and kept thawed until shipped to BGI-Shenzhen. Stool DNA was extracted using MagPure stool DNA KF kit B (no. MD5115-02B). DNA concentrations were estimated using Qubit (Invitrogen). The DNA library was constructed with 200ng DNA as input. Shotgun metagenomic sequencing was then performed on the BGI-seq platform^41^ to generate at least 10 million paired-end reads (length 100 bp) each sample.

#### Fudan cohort

For the Fudan cohort, sequencing data was downloaded from the NIH National Center for Biotechnology Information Sequence Read Archive (SRA) with BioProject ID PRJNA763023. In total, there were 200 metagenomes consisted of four individual groups (50 samples in each group): old control (oControl), old-onset colorectal cancer (oCRC), young control (yControl) and young-onset colorectal cancer (yCRC). The age cutoff for old and young-onset CRC was 50 years old. Details of the cohort can be found in Yang *et al*^18^.

#### Public cohort

The public cohort comprised samples from eight different studies^9–16^. Instead of downloading original sequencing reads, we obtained the processed MetaPhlAn 3 taxonomic and Humann 3 functional profiles from the supplementary table in Beghini et al^17^. There were 600 CRC patients and 662 controls. The number of samples in each cohort stratified by age was provided in **Supplementary Table 1**.

### Metagenomic sequencing data processing

The quality control of sequencing reads was done according to the metapi workflow (https://github.com/ohmeta/metapi/). Sequences with average quality score below Q30 (0.001% error rate) were removed. Adapter sequences and low-quality tails were trimmed. Only sequences with a length of 70bp or more were retained. High-quality sequences were then aligned to the human reference genome build hg38 with bowtie2^42^. Sequences mapped to the human genome under *–very-sensitive* mode were removed, and the resulting sequences were used in downstream analysis. **Supplementary Table 9** provides a summary of each quality control step. Since 449 out of 460 samples (97.6%) had >10 million paired-end reads for downstream analysis, we did not subsample the sequences to the lowest sequencing depth to avoid data loss.

To ensure the comparability of our analysis with several large CRC microbiome studies, we calculated the taxonomic and functional abundance profiles using MetaPhlAn 3 and HUMAnN 3^17^. Default parameters were used and the smallest relative abundance value was set to 1 × 10^-5^.

We also applied the same quality control and taxonomic analysis to the 200 metagenomes of Fudan cohort.

### Alpha and beta diversity

We assessed the microbiome alpha-diversity using Shannon index and the observed number of species, and beta-diversity using Bray-Curtis distance. The alpha-diversity was only evaluated on the species abundance profile, while the beta-diversity was evaluated on both the species and pathway abundance profiles. R package *vegan* was used for these calculations.

In our Guangzhou cohort, the Shannon index did not show a correlation with sequencing depth (rho= -0.027, P=0.56). We used permutation multivariate analysis of variance (PERMANOVA) based on the Bray-Curtis distance matrix to estimate the variance explained by age, sex, body mass index (BMI), smoking, tumor location, and stage in the Guangzhou cohort. We also used PERMANOVA to evaluate the disease and batch effect when integrating Guangzhou and Fudan cohorts. For each experiment, the number of permutations was set to 10,000.

### Identification of differential taxa and microbial pathway

To evaluate the correlation between the abundance of each species and age, we used Spearman correlation with and without correction for covariates. We corrected for covariates by first applying the central log-ratio (CLR) transformation to the relative abundance profile^43^, followed by fitting a linear regression model with sex, BMI, tumor location and stage, family history of CRC and smoking as covariates. We then calculated the Spearman correlations between the residuals and age. This analysis was performed on 372 individuals from the Guangzhou cohort who had no missing data for any covariate. As most of the previously reported CRC-associated microbes presented in >15% of the individuals in Guangzhou cohort, we only applied the testing to taxa with a prevalence rate of at least 15% to ensure the power of analysis.

We used Wilcoxon–Mann–Whitney test to identify microbial taxa and pathways with differential abundance between CRC patients and controls in the Fudan (50 oCRC, 50 oControl, 50 yCRC, and 50 yControl) and Guangzhou cohorts (293 oCRC and 167 yCRC). We conducted four different comparisons: (1) Fudan oCRC versus Fudan oConrol, (2) Fudan yCRC versus Fudan yControl, (3) Guangzhou oCRC versus Fudan oControl, and (4) Guangzhou yCRC versus Fudan yControl. We corrected *P* values for multiple hypotheses using Benjamini-Hochberg (FDR) procedure for all taxa in each comparison. This analysis was conducted in R.

### Strain-level analysis of *F. nucleatum*, *B. fragilis* and *E. coli*

We used two distinct methods, StrainPhlAn3^17^ and inStrain^28^, to explore the strain-level diversity of three well-known CRC-associated bacteria: *F. nucleatum* (*Fn*), *B. fragilis* (*Bf*) and *E. coli* (*Ec*). StrainphlAn3 relies on marker genes, while inStrain considers the genome-wide sequences. Our analysis focused on 460 Guangzhou samples, which had higher sequencing depth than Fudan and public samples.

For StrainPhlAn3, default parameters were applied, requiring a minimum of 20 marker genes per sample, each marker gene found in at least 80% samples. *Fn* was found in 63 (14%) samples based on 36 marker genes. *Bf* was found in 334 (73%) samples based on 46 marker genes. *Ec* was found in 317 (69%) samples based on 24 marker genes.

For inStrain analysis, we built a comprehensive reference genome from *de novo* assembly and NCBI RefSeq genomes. Using the metapi pipeline, which employed megahit v1.2.9^44^ for genome assembly, metabat2^45^ for binning, and bowtie2 for alignment, we obtained 37,441 metagenome-assembled genomes (MAGs) with a minimum length of 200 kb. We then used GTDB toolkit v2.1.0^46^ to annotate these MAGs referring to GTDB release207^47^ with default parameters. For *Fn*, only 6 samples had MAGs annotated to *Fusobacterium nucleatum_J* (RefSeq assembly GCF_008633215.1, NCBI strain identifier 13-08-02), while 11 samples had MAGs annotated to *Fusobacterium animalis* (*Fa*, RefSeq assembly GCF_000158275.2, strain identifier 7_1). *Fa* was also known as *Fn subsp. animalis*. For *Bf*, 261 samples had MAGs annotated to RefSeq assembly GCF_000025985.1 (NCBI strain NCTC 9343), and 60 samples had MAGs annotated to RefSeq assembly GCF_002849695.1 (NCBI strain Q1F2). For *Ec*, 285 samples had MAGs annotated to RefSeq assembly GCF_003697165.2 (NCBI strain ATCC 11775). As a sanity check, samples with MAGs annotated to *Fn*, *Bf* and *Ec* strains were recovered in the corresponding StrainPhlAn3 analysis. We combined 6,356 high quality MAGs (>90% completeness and <5% contamination, according to the definition in Bowers et al.^48^) and 5 RefSeq genomes (mentioned above) together and used dRep^49^ to select a unique set of reference genomes. After dereplication, 3,084 genomes remained, including the 5 RefSeq genomes. Subsequently, reads from 460 Guangzhou samples were aligned to the dereplicated genomes using bowtie2, and the inStrain analysis was applied to the 5 RefSeq genomes.

Given the varied abundances of different taxa, we used taxon-specific parameters for the interpretation of inStrain results. For *Fn* and *Fa*, we used breadth >0.1 and coverage >0.1(meaning that at least 10% of the reference genome was mapped and the average genome-wide mapping frequency is at least 0.1); for *Bf*, breadth >0.5 and coverage >1.0; for *Ec*, breadth >0.1 and coverage >0.2. These criteria retained 50 samples for *Fn* (including 5 with MAGs annotated to RefSeq assembly GCF_008633215.1 and 22 supported by StrainPhlAn3) and 84 samples for *Fa* (including 11 with MAGs annotated to RefSeq assembly GCF_000158275.2). For *Ec*, 328 samples met the criteria, with 299 (91%) supported by StrainPhlAn3.

For *Bf*, 275 samples remained for strain NCTC9343 (271 samples supported by StrainPhlAn3) and 67 samples for strain Q1F2 (66 samples supported by StrainPhlAn3). Of note, strain Q1F2 was not covered in StrainPhlAn3 due to the absence of marker genes for this taxon. We inspected the gene marker-based phylogenetic tree of *Bf.* There were two distinct clusters, one was predominantly composed of samples with MAGs annotated to strain NCTC9343, and the other was dominated by samples with MAGs annotated to strain Q1F2. Consequently, we manually divided the 334 StrainPhlAn3 samples into two clusters. Cluster 1 (related to NCTC9343) comprised 267 samples, all supported by inStrain analysis. Cluster 2 (related to Q1F2) consisted of 67 samples, with 61 supported by inStrain analysis.

### Known CRC associated microbial taxa

Known CRC associated microbial taxa were obtained from the gutMDisorder^24^ (http://bio-annotation.cn/gutMDisorder) database with parameters: *Species=Human, Condition=Colorectal Neoplasms (distal cancer), Sequencing Technology=Whole metagenomic sequencing* and *Taxonomy Rank=species*. The resulting list of 148 records covering 118 species was downloaded in March 2023 and shown in **Supplementary Table 10**.

### CRC-associated genes

We obtained reference sequences of CRC-associated genes from Wirbel *et al*.^14^. Specifically, representative sequences for *fadA*, *bft*, the *pks* genomic island and the *bai* operon were identified from integrated gene catalog (IGC)^50^ using gene-specific Hidden Markova Model. Subsequently, metagenomic sequencing reads were mapped to the IGC using bowtie2, and the reads count for each gene was calculated. The number of mapped reads for each gene was then normalized by dividing it by the total number of mapped reads in each sample. The resulting number can be interpreted as the number of reads per million mapped reads (RPM). The abundances of *fadA*, *bft*, the *pks* and *bai* were the sum of all their member genes. For the *cutC* gene, we used the output from Humman3 calculations for the gene family. The abundance of the *cutC* gene was determined as the sum of its member gene clusters.

### Microbiome-based classification

We evaluated the microbiome-based classification capabilities for oCRC and yCRC using two widely adopted algorithms: random forest and least absolute shrinkage and selection operator (LASSO) logistic regression. The random forest algorithm, known for its superior performance among various machine learning tools^51^, has been successfully employed in previous large-scale CRC microbiome studies^13,17^. The LASSO logistic regression has been used in a comprehensive meta-analysis study on CRC microbiome^14^. The assessment was conducted on two types of microbiome quantitative profiles: species and pathway-level relative abundances calculated by MetaPhlAn3 and HUMAnN3. To enhance the generalizability of our models, we filtered out features not presented in all studied cohorts. This resulted in a dataset comprising 199 taxa and 401 pathways that were consistently found in at least one sample in each of the Guangzhou, Fudan and public cohorts.

In the random forest task, we used the algorithm implemented in R package *mlr3*. Each training iteration consisted of an ensemble of 10,000 estimator trees, with the number of features per tree set to the square root of the total number of features. The impurity score was determined by Shannon entropy (‘*gini’*) to evaluate the quality of tree growing. Other parameters included no-maximum depth for the trees and one sample as the minimum amount for leaf node. To estimate the within-dataset prediction capability, ten-fold cross-validation was employed for Fudan oCRC and yCRC, and this process was repeated 100 times. The reported results represent an average over 100 validation folds. For cross-study prediction, the model was trained once on all the training samples and applied to the testing samples. These experiments were conducted using the output values from MetaPhlAn3 and HUMAnN3, as the random forest algorithm is robust to data normalization.

In the LASSO logistic regression task, the relative abundances were log_10_-transformed and standardized as z-scores. Zeros in the species and pathway abundance table were replaced by a pseudo-count of 1 × 10^-6^ and 1 × 10^-8^ before the log_10_-transformation. During the training phase, a ten-fold cross-validation strategy was employed to tune the lambda parameter (regulation strength). The lambda parameter for each model was selected to maximize the area under the precision-recall curve. We repeated the ten-fold cross-validation 100 times for models trained exclusively on Fudan data and 10 times for models trained on public data (with or without Fudan and Guangzhou data). In the prediction phase, the prediction evaluation was averaged across all models. For models trained on Fudan data, a nested feature selection step was implemented, and the models with the best performance were reported. All these experiments were conducted in R with package SIAMCAT^52^.

### Statistical analysis

To access the correlation between age and metadata, Guangzhou CRC patients were grouped into six age categories: [20,30), [30,40), [40,50), [50,60), [60,79) and >70. Subsequently, the distribution of sex, tumor stage and location, family history of CRC, smoking status, and tumor molecular characteristics (mismatch repair, MSH2, MSH6, MLH1, PMS2, HER2 and BRAF status) across these age groups was examined using the chi-square test. The Fisher test was employed if any count was less than 5. Furthermore, patients were divided into two groups based on age, specifically, old-onset (age≥50) and young-onset (age<50), and the same tests were repeated. For body max index (BMI), the means in oCRC and yCRC were compared using the Wilcoxon–Mann– Whitney test. The association between BMI and age in all patients was measured using Pearson correlation. The R functions *chisq.test()*, *fisher.test()*, *wilcox.test()* and *cor.test()* were used for these analyses.

## Data availability

The data that support the findings of this study have been deposited into CNGB Sequence Archive (CNSA)^53^ of China National GeneBank DataBase (CNGBdb)^54^ with accession number CNP0004314. This archive contains the fastq sequences, *de novo* assembly, and binning results. A copy of data has also been deposited into Genome Sequence Archive (GSA, https://ngdc.cncb.ac.cn/gsa/) with submission number of HRA004617. The microbiome data and associated codes used in microbiome-based classification tasks are available on GitHub repository https://github.com/Owen-haha/CRCmicrobiome.

## Acknowledgements

The study was supported by the National Natural Science Foundation of China [82073159, 82002467], the Natural Science Foundation of Guangdong [2022A1515012296], Beijing Xisike Clinical Oncology Research Foundation [Y-tongshu2021/ms-0175], Guangdong Genomics Data Center [2021B1212100001]. We thank all the participating patients for their generous and volunteer support of this study. We thank the Fudan research team for their efforts to make the data publicly available. We thank the China National GeneBank (CNGB) Shenzhen for DNA extraction, library construction, and sequencing. We gratefully thank our BGI colleagues Jie Zhu, Yanmei Ju, Weiting Liang, Liu Tian, Xiaoqian Lin and Tongyuan Hu for their technical supports in bioinformatics.

## Figure legends

**Supplementary Figure S1.**
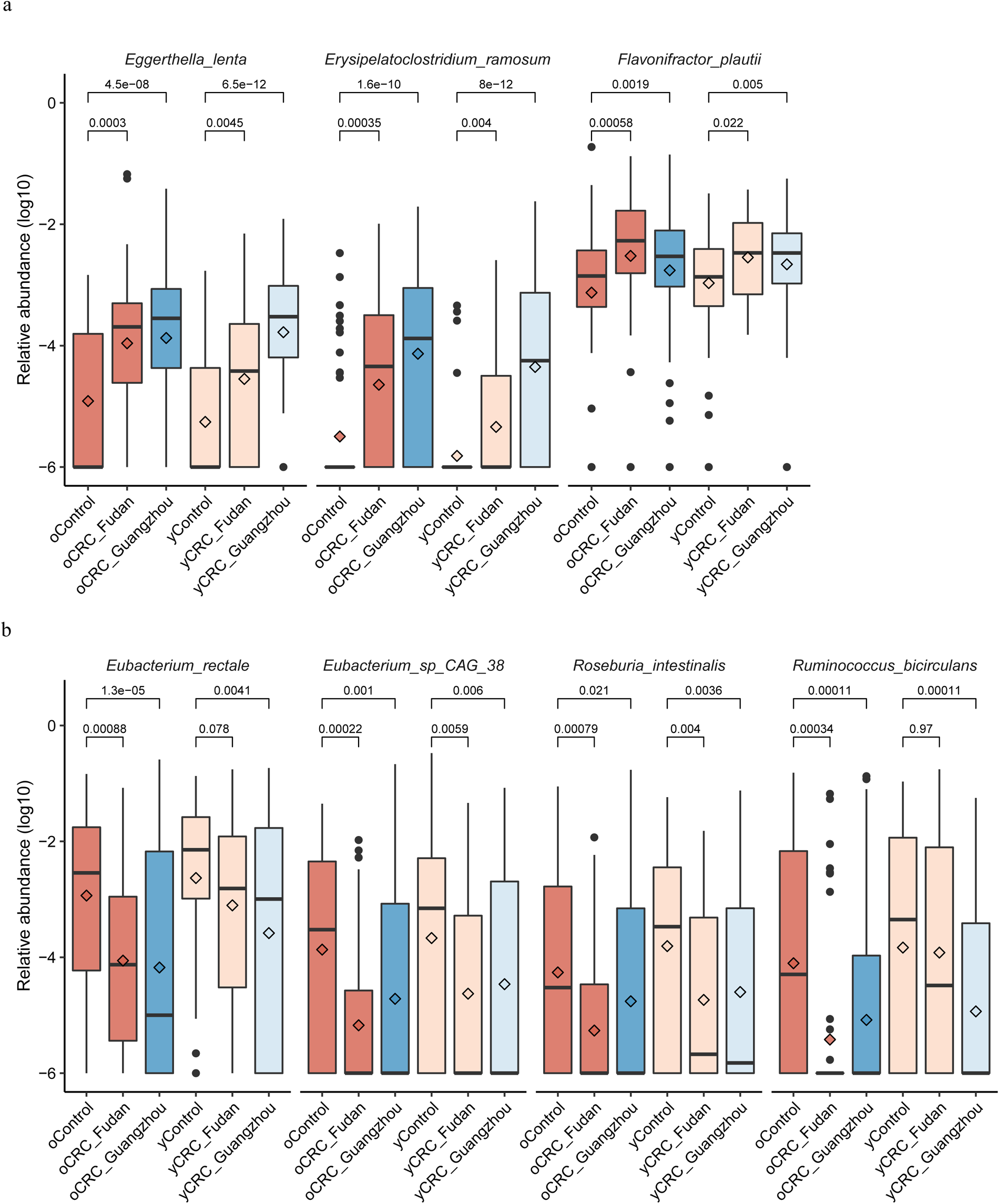
Abundance distribution of taxa significantly differentiated in oCRC but not in yCRC. Top: Three microbial taxa enriched in oCRC groups at FDR adjusted P<0.05. These taxa were also increased in yCRC groups but with an FDR adjusted P value above 0.05. Bottom: Four microbial taxa depleted in oCRC groups (FDR adjusted P<0.05). These taxa were also depleted in yCRC groups but with an FDR adjusted P value above 0.05. P values on the top were calculated by Wilcoxon rank-sum test (no FDR adjustment). The relative abundance is in log10 scale and zeros were replaced by a small value. The boxplots show the median (thick line), interquartile range (box limits), 1.5× the interquartile range span (whiskers) and outliers (dots). Diamond shape indicates the mean abundance.

**Supplementary Figure S2.**
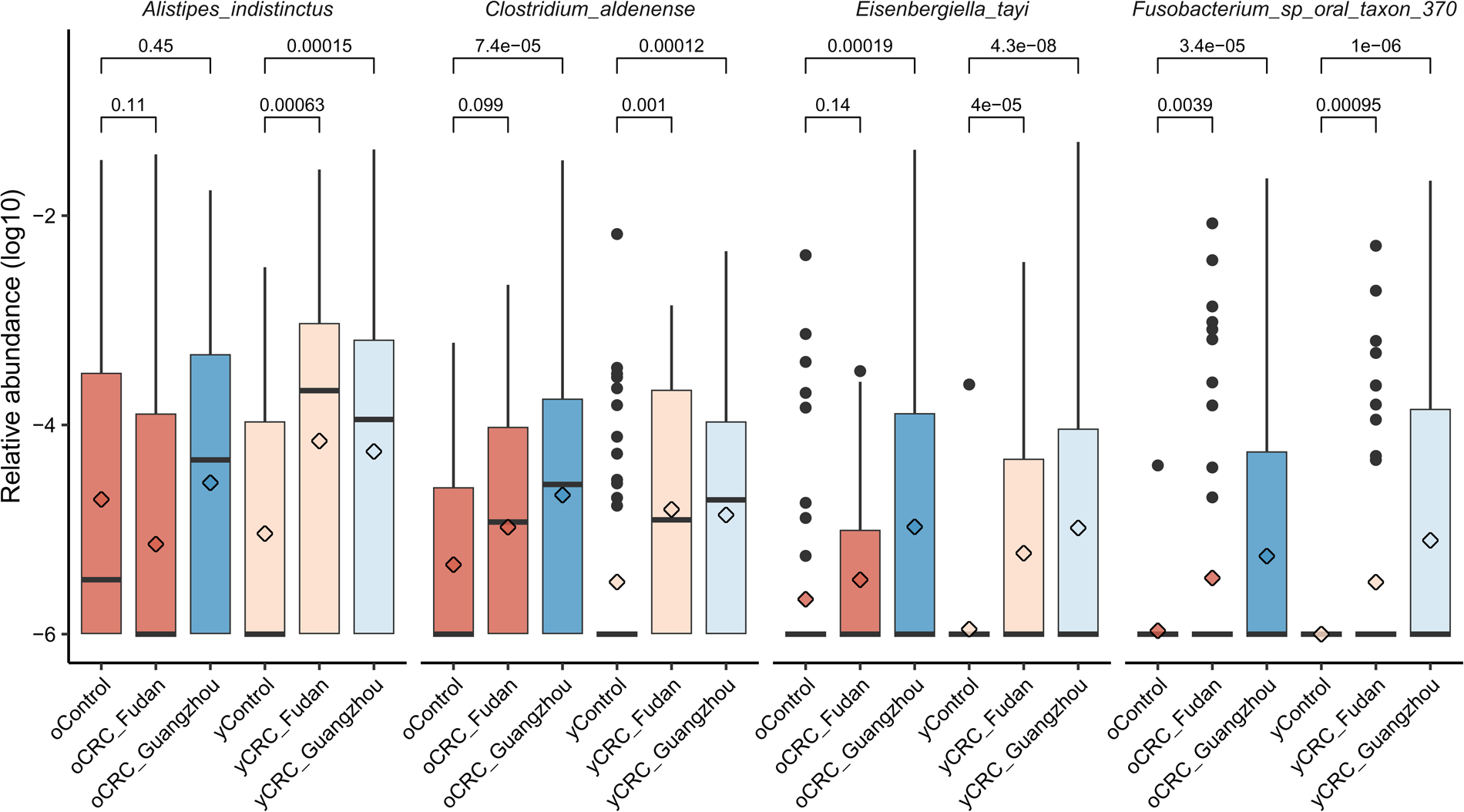
Abundance distribution of taxa significantly differentiated in yCRC but not in oCRC. Four microbial taxa enriched in yCRC groups at FDR adjusted P<0.05. Three of them were also increased in oCRC groups but with an FDR adjusted P value above 0.05. P values on the top were calculated by Wilcoxon rank-sum test (no FDR adjustment). The relative abundance is in log10 scale and zeros were replaced by a small value. The meaning of boxplot is same to the description in **Figure S1**.

**Supplementary Figure S3.**
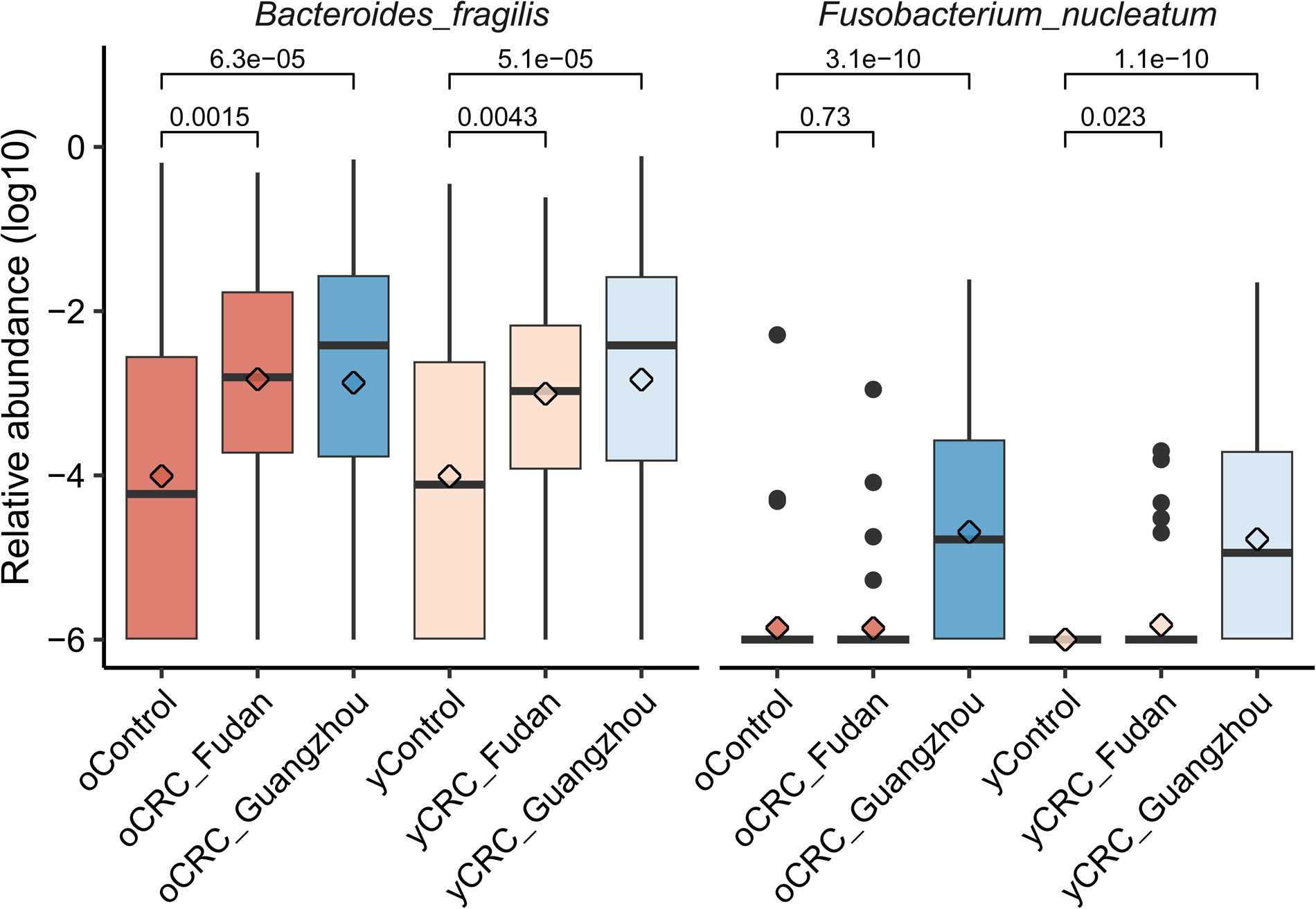
Abundance distribution of the well-known CRC-enriched taxa *Bacteroides fragilis* and *Fusobacterium nucleatum* with potential carcinogenesis mechanism. The relative abundance is in log10 scale and zeros were replaced by a small value. P values on the top were calculated by Wilcoxon rank-sum test (no FDR adjustment).

**Figure S4:**
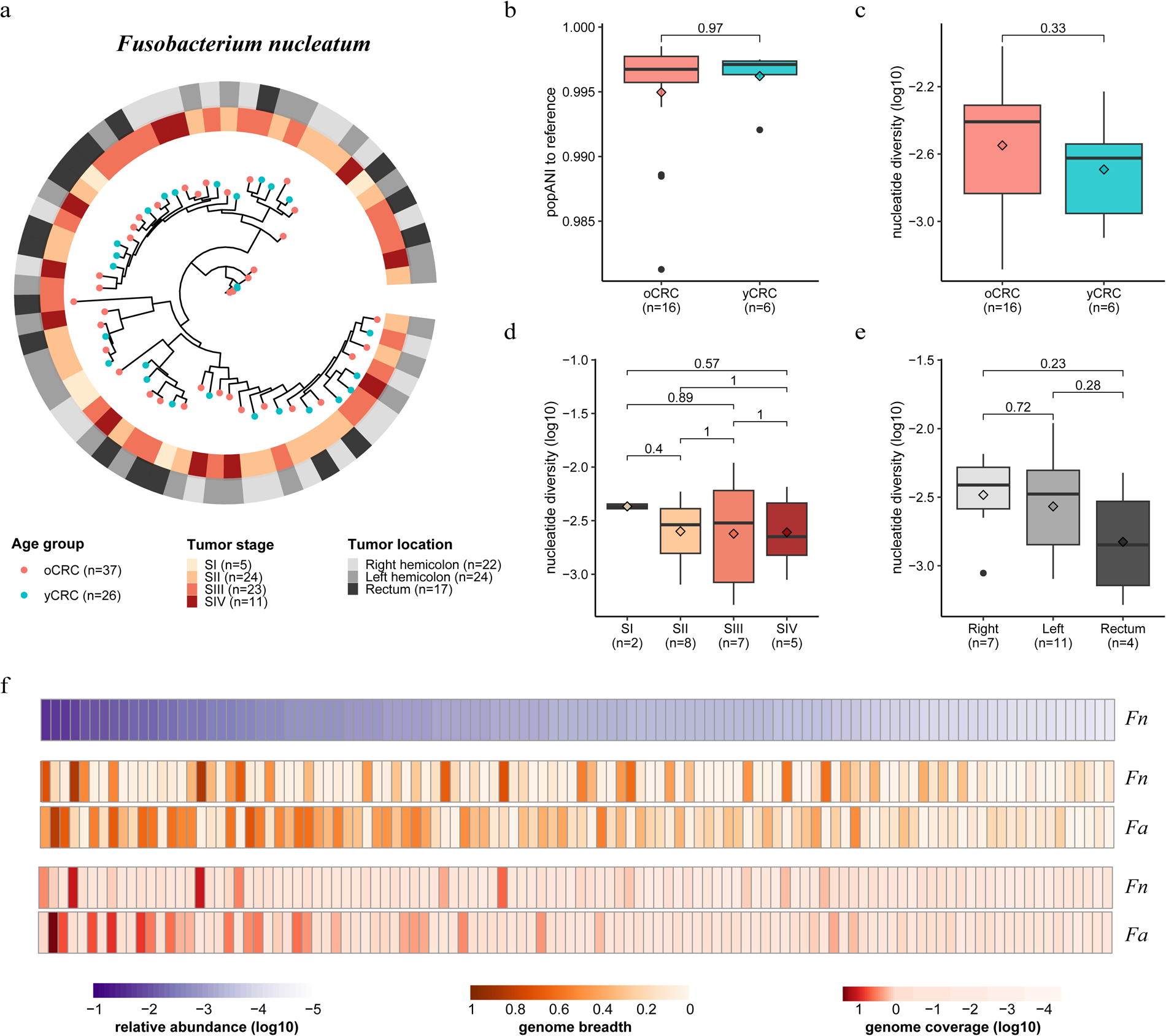
Phylogenetic and genomic analysis of *Fusobacterium nucleatum* in CRC. (**a**) Phylogenetic tree of *F. nucleatum* constructed based on 36 marker genes. Tips are samples and colored by age group. Rings outside the tree indicate tumor stage and location. Only samples with reads mapped to at least 20 marker genes are displayed. (**b**) Population average nucleotide identity (popANI) values to reference genome (RefSeq GCF_008633215.1). (**c**), (**d**) and (**e**) are values of genome-wide nucleotide diversity stratified by age, tumor stage and location. Only panel (**a**) samples with reads mapped to the reference genome reaching a genome-wide breadth >0.1 and coverage >0.1 are shown. (**f**) Heatmap showing the abundance, genome-wide breadth, and coverage of *F. nucleatum* (*Fn*) and *F. animalis* (*Fa*). Only 110 samples which had genome breadth >0.1 and coverage >0.1 for *Fn* (RefSeq GCF_008633215.1) or *Fa* (RefSeq GCF_000158275.2) reference genomes are included. Samples are sorted by the relative abundance of *Fn*, calculated by MetaPhan3. Genome breadth and coverage were determined using inStrain. Boxplot conventions are consistent with the description in **Figure S1**.

**Figure S5:**
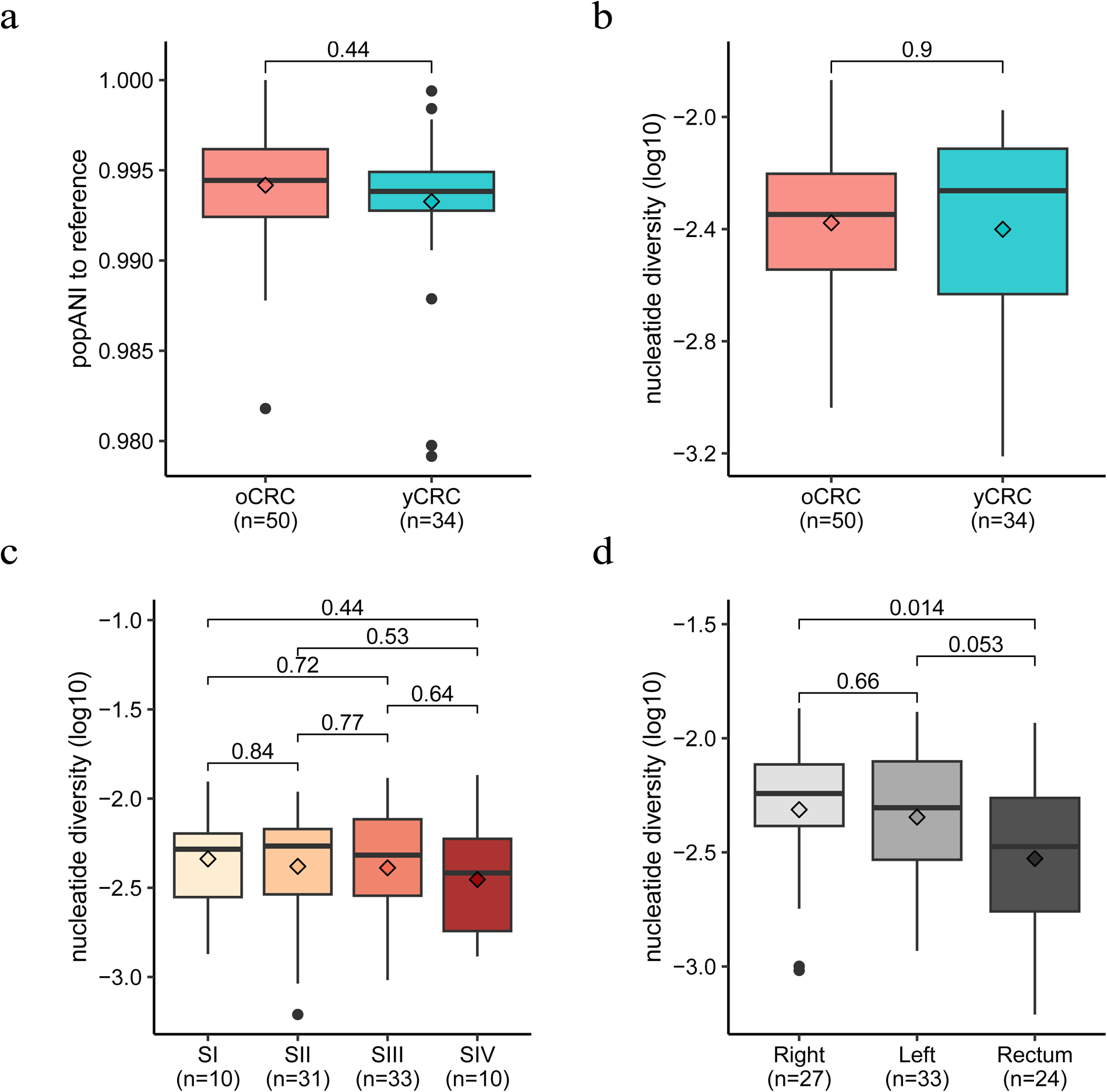
Genomic analysis of *Fusobacterium animalis* in CRC. (**a**) Population average nucleotide identity (popANI) values to reference genome (RefSeq GCF_008633215.1). (**b**), (**c**) and (**d**) are values of genome-wide nucleotide diversity stratified by age, tumor stage and location. Only panel samples with reads mapped to the reference genome reaching a genome-wide breadth >0.1 and coverage >0.1 were shown. Boxplot conventions are consistent with the description in **Figure S1**.

**Figure S6:**
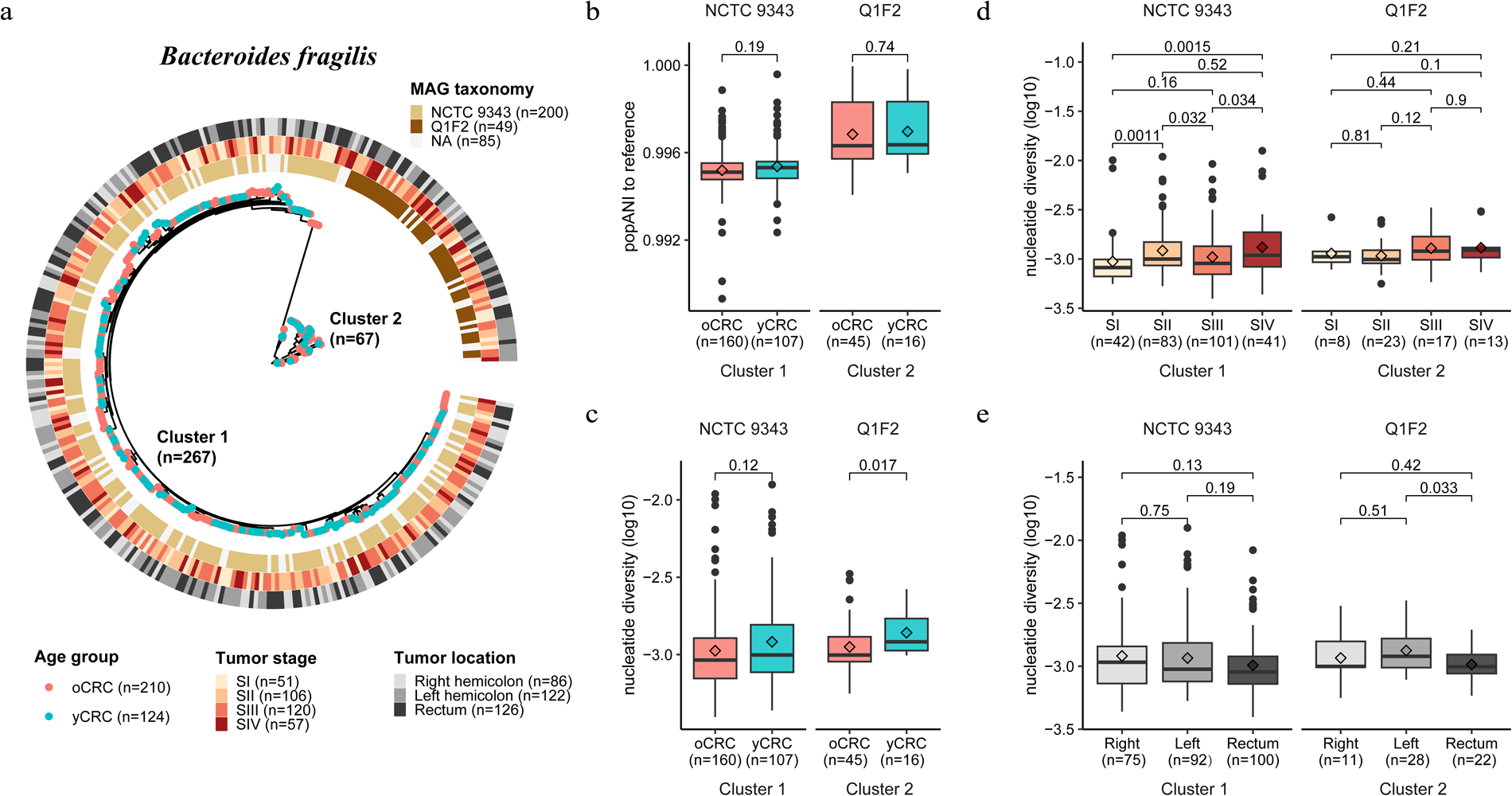
Phylogenetic and genomic analysis of *Bacteroides fragilis* in CRC. (**a**) Phylogenetic tree of *B. fragilis* constructed based on 46 marker genes. Two distinct strain clusters are shown, and we named them as cluster 1 (dominated by samples with metagenome-assembled genomes (MAGs) annotated as strain NCTC 9343) and cluster 2 (dominated by samples with MAGs annotated as strain Q1F2). Tips are samples and colored by age group. Rings outside the tree indicate MAG taxonomy, tumor stage and location. Only samples with reads mapped to at least 20 marker genes are displayed. (**b**) Population average nucleotide identity (popANI) values to reference genomes of strain NCTC 9343 (RefSeq GCF_000025985.1) and Q1F2 (RefSeq GCF_002849695.1). (**c**), (**d**) and (**e**) are values of genome-wide nucleotide diversity stratified by strain cluster, age, tumor stage and location. Only panel (**a**) samples with reads mapped to the reference genome reaching a genome-wide breadth >0.5 and coverage >1 were shown. Boxplot conventions are consistent with the description in **Figure S1**.

**Figure S7:**
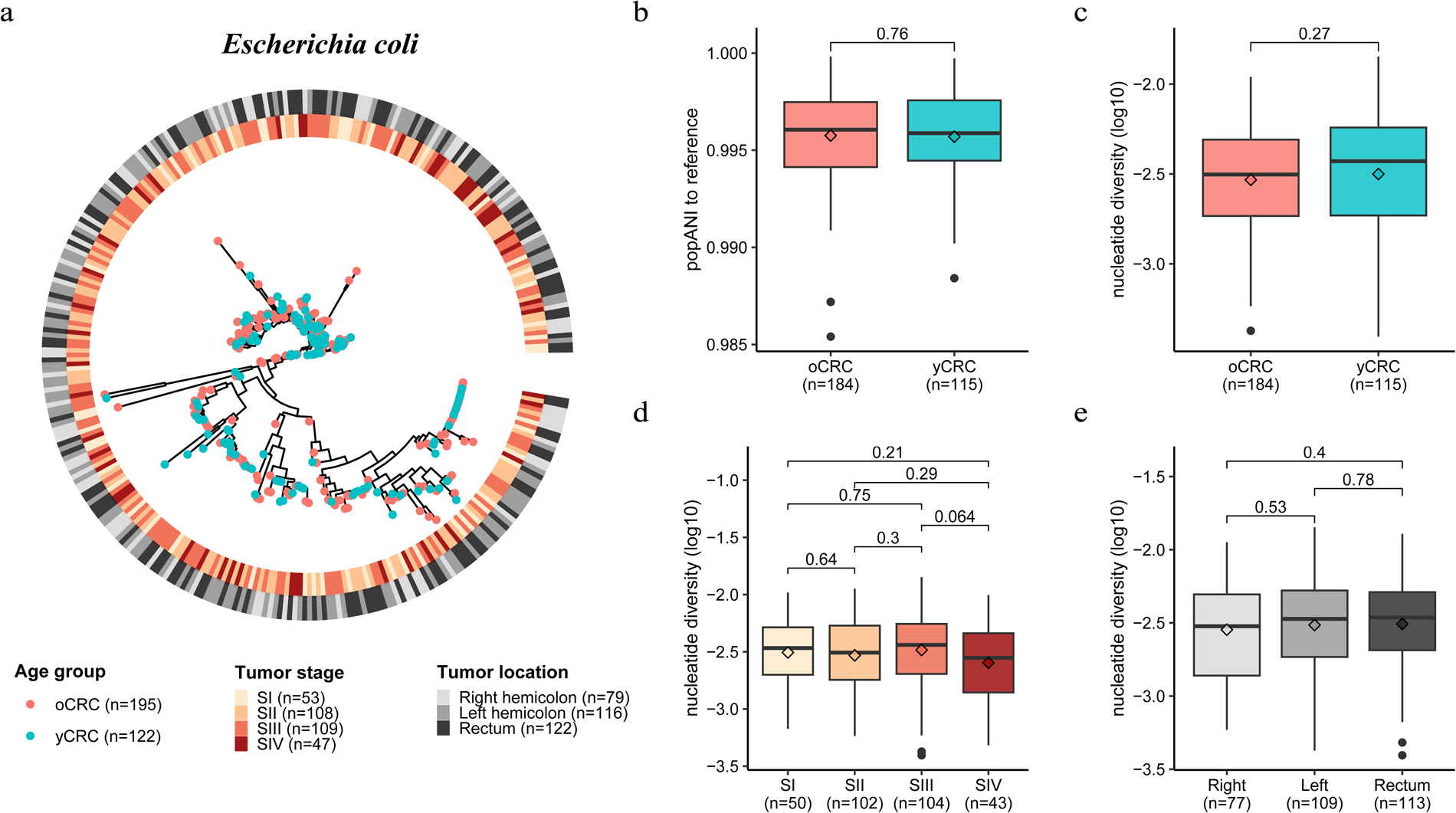
Phylogenetic and genomic analysis of *Escherichia coli* strain in CRC. (**a**) Phylogenetic tree of *E. coli* constructed based on 24 marker genes. Tips are samples and colored by age group. Rings outside the tree indicate tumor stage and location. Only samples with reads mapped to at least 20 marker genes are displayed. (**b**) Population average nucleotide identity (popANI) values to reference genome (RefSeq GCF_003697165.2). (**c**), (**d**) and (**e**) are values of genome-wide nucleotide diversity stratified by age, tumor stage and location. Only panel (**a**) samples with reads mapped to the reference genome reaching a genome-wide breadth >0.1 and coverage >0.2 were shown. Boxplot conventions are consistent with the description in **Figure S1**.

**Supplementary Figure S8.**
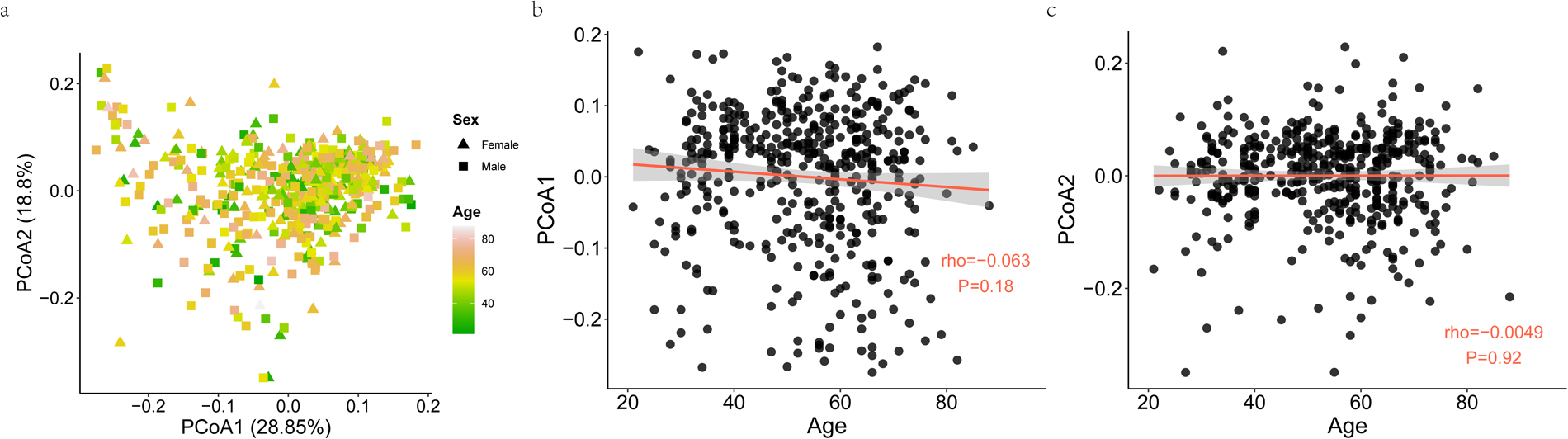
Overall distribution of the Guangzhou samples based on microbial pathway profile. (**a**) Two-dimension scatter plot shows the overall pattern of samples. Principle coordinate analysis (PCoA) was performed based on the Bray-Curtis distance. Each point represents one sample and color scale indicates age. Samples from female and male patients are in triangles and squares, respectively. Scatterplot of relationship between age and PCoA axis 1 (**b**) and PCoA axis 2 (**c**). The solid red line was fitted by smooth function in R and the grey area is the 95% confidence interval.

**Supplementary Figure S9.**
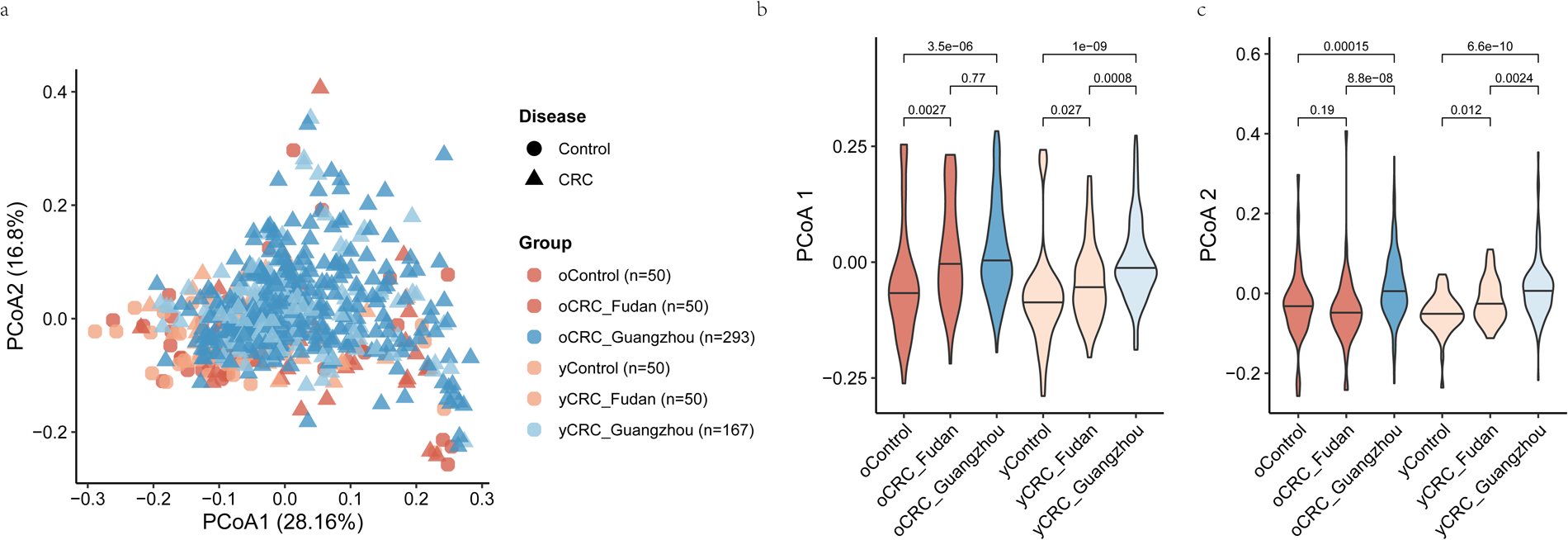
Overall distribution of the Guangzhou and Fudan samples based on microbial pathway profile. (**a**) Two-dimension scatter plot shows the overall distribution of Fudan and Guangzhou samples. Principle coordinate analysis (PCoA) was performed based on the Bray-Curtis distance. Each point represents one sample. Samples from Fudan and Guangzhou cohorts are in red and blue, respectively. Circles are control samples, while triangles are CRC samples. Violin plots show values of PCoA axis 1 (**b**), PCoA axis 2 (**c**). P values on the top were calculated by Wilcoxon rank-sum test. The thick horizon line indicates the 50% percentile.

**Supplementary Figure S10.**
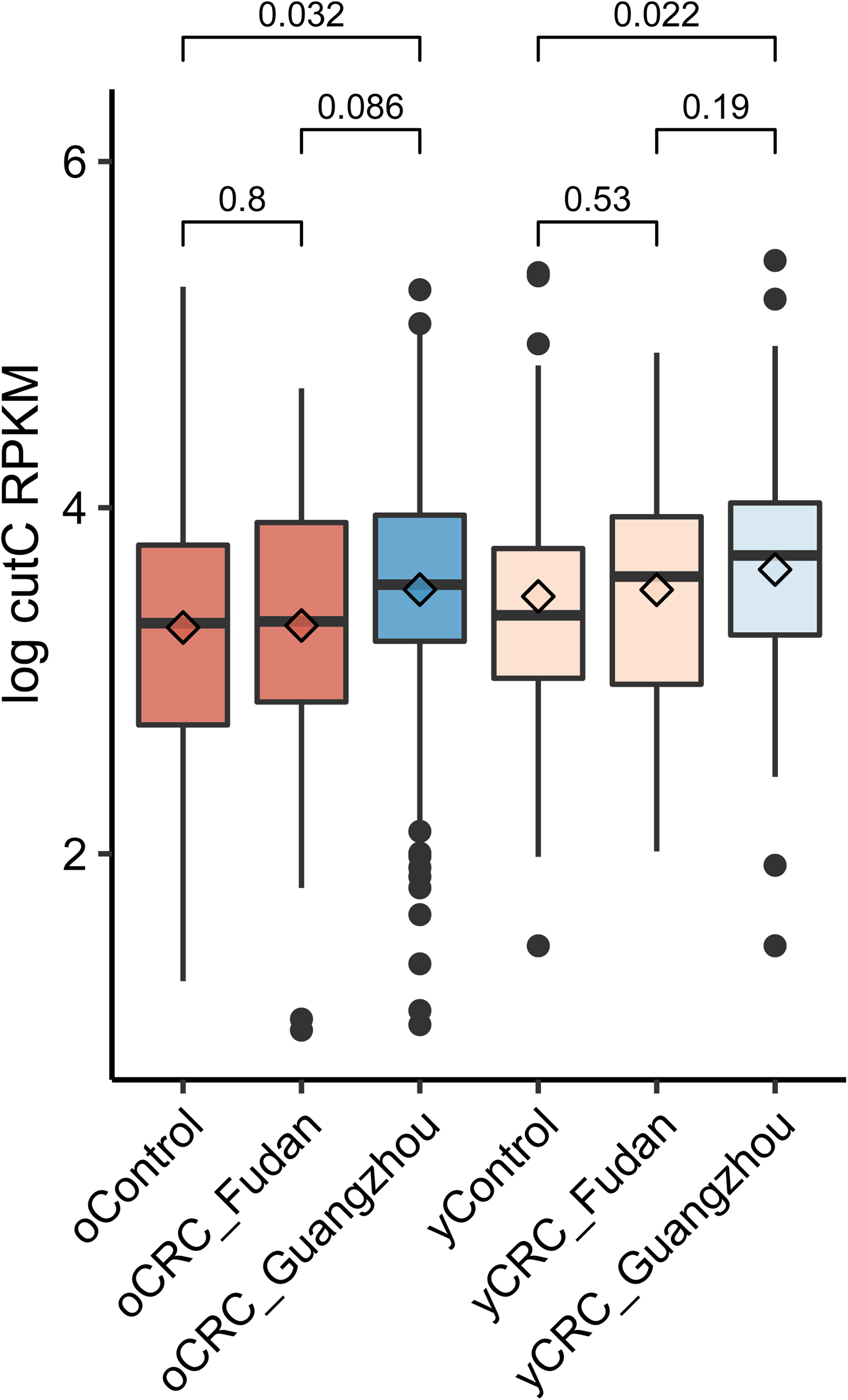
Abundance distribution of the well-known CRC-enriched microbial *cutC* gene. The y-axis shows the number of mapped reads per kilobase per million reads (RPKM) in log scale. P values on the top were calculated by Wilcoxon rank-sum test. Boxplot conventions are consistent with the description in **Figure S1**.

**Supplementary Figure S11.**
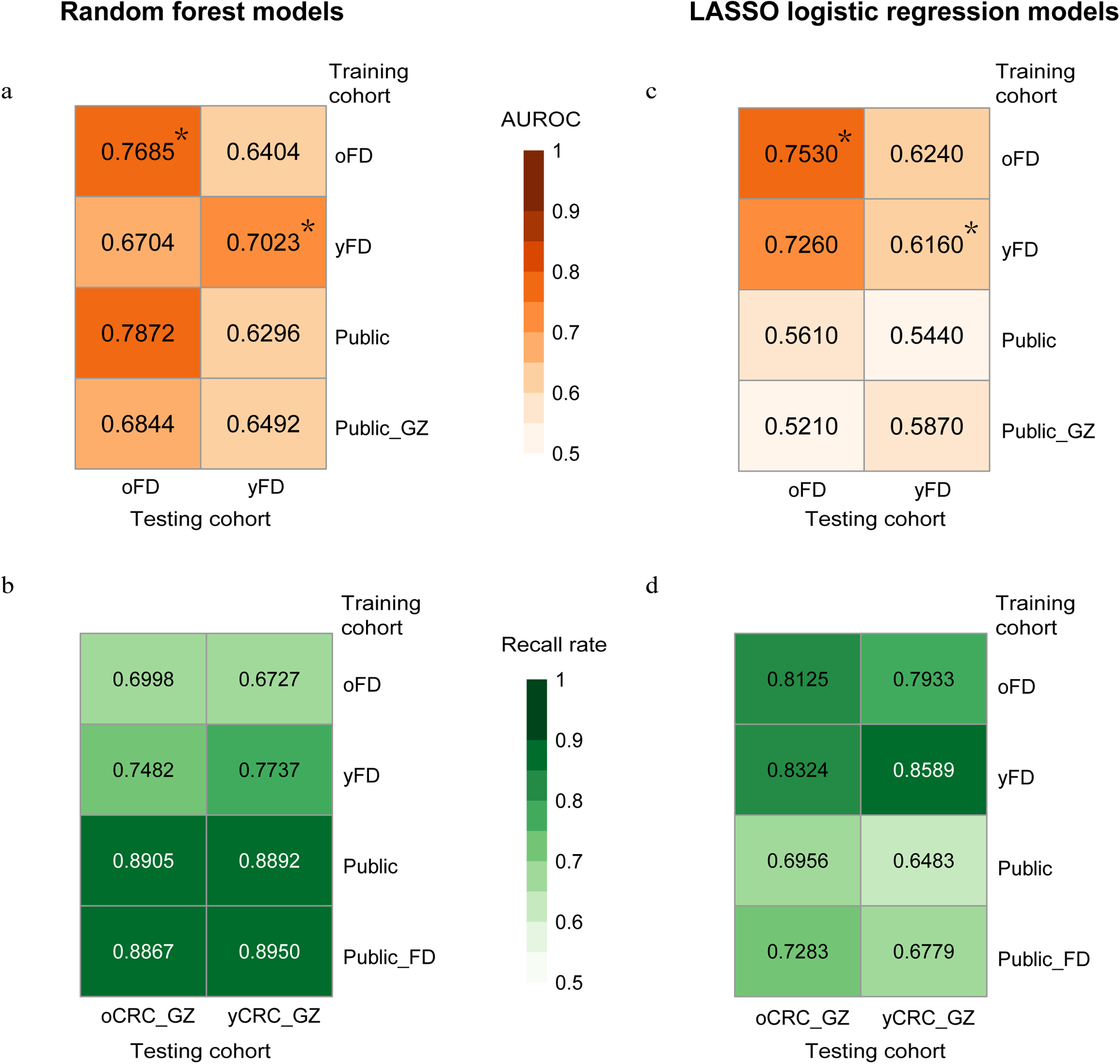
Prediction performance of microbial pathway-based classification models. Prediction performance on oCRC and yCRC in Fudan and Guangzhou cohorts for models trained on pathway-level abundances from different datasets. Models were trained on two different methods: random forest and LASSO logistic regression. The numbers are the area under receiver operator curve (AUROC) for (**a** & **c**), and recall rate for (**b** & **d**). Asterisks denote values averaged over 100 times ten-fold cross-validation. Abbreviations: oFD means the 100 metagenomes of Fudan oCRC and oControl; yFD means the 100 metagenomes of Fudan yCRC and yControl; Public means 1,262 public metagenomes; Public_GZ means 1,262 public metagenomes plus 460 Guangzhou metagenomes; Public_FD means 1,262 public metagenomes plus 200 Fudan metagenomes.

